# Non-Blood Liquid Biopsy of Nipple Discharge to Decode Tumor-Derived CNV Signatures for Breast Cancer Detection and Prognostication

**DOI:** 10.64898/2026.11.24.26365856

**Authors:** Yalun Li, Pengming Pu, Hengyi Xu, Ruijie Zhou, Pengrui Tang, Wen Wen, Guangdong Qiao, Yeqing Zhou, Ge Gao, Yuchen Liu, Yansong Huang, Zizhao Guo, Tongxuan Shang, Lin Cong, Jiahui Xiang, Jiang Wu, Dongxu Ma, Hezhi Fang, Jiaqi Liu

## Abstract

Accurate, noninvasive diagnosis of malignant nipple discharge remains a longstanding clinical challenge. This study evaluated somatic copy-number variations (CNVs) in nipple discharge DNA (ndDNA) as high-fidelity diagnostic biomarkers and explored their cellular origins and shedding mechanisms. We derived a 20-feature CNV panel from large-scale genomic datasets and validated its diagnostic potential. Integrative single-cell analysis of 50 patients revealed that these features predominantly arose in ancestral states across molecular subtypes and exhibited functionally relevant co-occurrence patterns. In clinical cohorts, ndDNA-based detection achieved 74.4% sensitivity and 94.4% specificity (AUC = 0.863), significantly outperforming plasma cell-free DNA (AUC = 0.517; p < 0.0001). Paired single-cell RNA sequencing confirmed high concordance between ndDNA profiles and malignant epithelial cells. High-fidelity epithelial subclusters displayed active estrogen signaling and distinct immune interactions, and a derived Fidelity Score independently predicted favorable prognosis in two external survival cohorts. Our findings establish nipple discharge as a tumor-proximal liquid biopsy that preserves early tumor genomic signatures with minimal dilution, advancing noninvasive diagnostic and prognostic strategies.

**Graphic Abstract:** 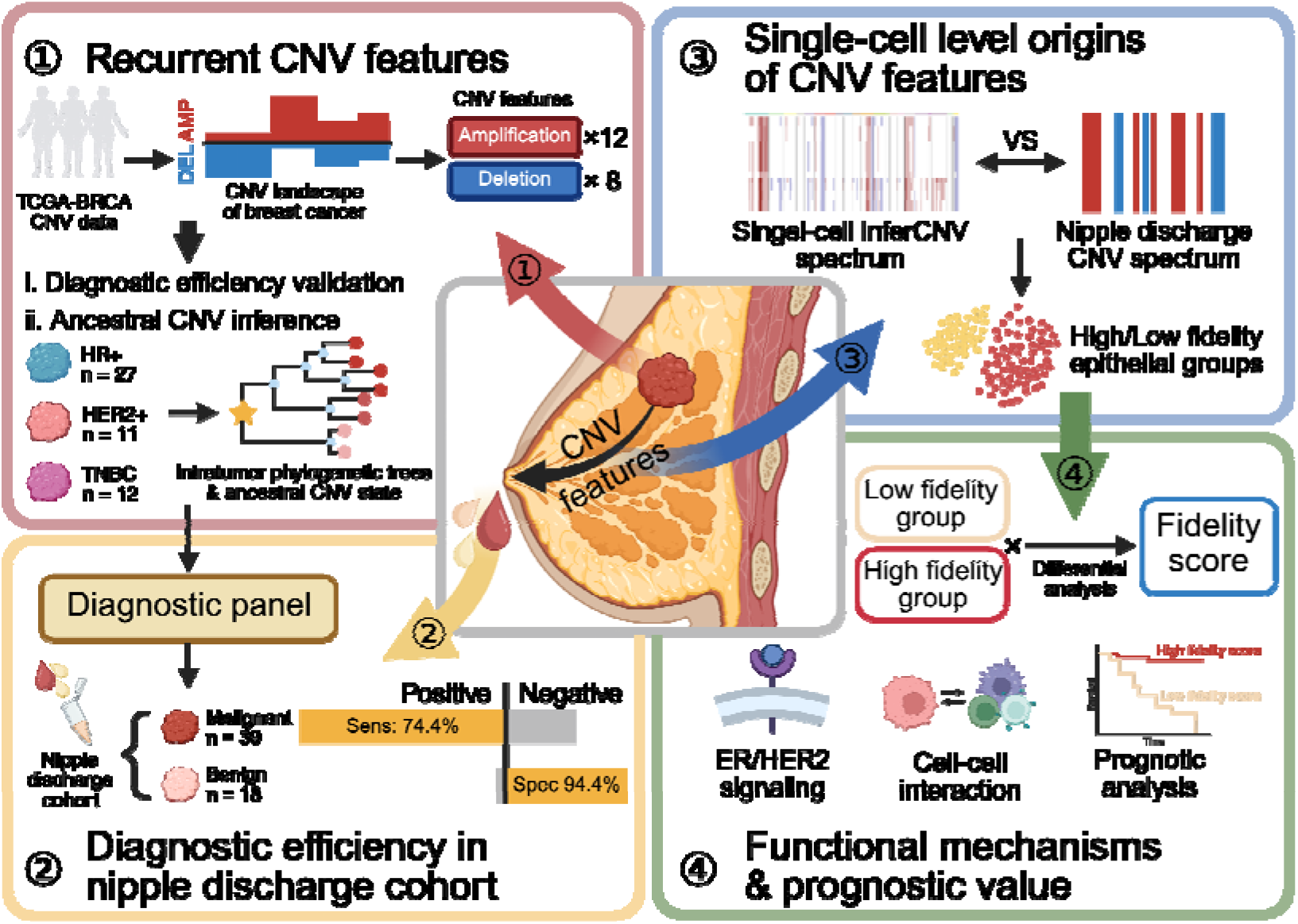

## Background

Breast cancer remains the most prevalent malignancy and one of the leading causes of cancer death among women worldwide[1]. Accurate assessment of symptoms indicative of malignancy is critical for improving outcomes[2]. Palpable breast masses benefit from a well-established, multimodal diagnostic workflow that includes mammography, ultrasound, and magnetic resonance imaging[3]. In contrast, evaluating nipple discharge remains a pressing clinical dilemma, as it is a common symptom affecting ∼80% of women during reproductive years[4, 5] while serving as the presenting complaint in only 4% of breast cancer patients[6, 7]. Current diagnostic methods for nipple discharge, such as ductoscopy and cytology, are hampered by insufficient sensitivity (50% and 62%, respectively) and specificity (83% and 71%, respectively)[8, 9]. These limitations directly lead to unnecessary invasive procedures in benign cases and delayed diagnosis in early malignant cases, creating a longstanding unmet clinical need for reliable, noninvasive alternatives.

Recent advances in liquid biopsy have expanded beyond blood to include biofluids that are in closer proximity to tumor sites, offering potentially richer diagnostic information[10–12]. For example, tumor-derived DNA in bladder cancer is consistently more detectable in urine than in paired plasma[13]. Furthermore, the copy-number variation (CNV) profiling of urine cell-free DNA (cfDNA) using low-pass whole-genome sequencing (LP-WGS) outperforms plasma-based analysis in detecting minimal residual disease and predicting survival in bladder cancer patients[14]. Similarly, cerebrospinal fluid circulating tumor DNA (ctDNA) more accurately reflects the genomic profile of central nervous system metastases than plasma ctDNA, thereby improving risk stratification and therapy guidance[15]. Malignant ascites-derived tumor cells are directly descended from peritoneal metastases[16]. Moreover, as evidenced by a recent breast cancer study, the proximity of non-blood fluids (ascites, cerebrospinal fluid, pericardial fluid, pleural fluid, saliva, and urine) to metastatic tumor sites enables them to frequently capture clinically relevant genomic alterations not observed in blood[17]. Given that nipple discharge originates directly from the mammary ductal system—the anatomical site of breast tumor initiation and progression—it is inherently poised to serve as an ideal liquid biopsy substrate. However, unlike other non-blood biofluids, its potential as a genomic diagnostic source for breast cancer remains largely unexplored.

***Genome instability*** is a fundamental hallmark of cancer, and CNVs are considered early oncogenic drivers[18]. Their stability, ubiquity, and tumor specificity make CNVs ideal biomarkers detectable across bodily fluids[15, 19–21]. A recent phenome-wide analysis of 470,727 whole-genome sequences has also demonstrated that CNVs exhibit large effect sizes and robust associations with diverse clinical phenotypes, underscoring their utility as high-impact genomic markers[22]. However, the comprehensive landscape of recurrent CNVs in breast cancer remains incomplete, which consequently obscures their diagnostic potential. Furthermore, the diagnostic efficacy of using CNVs as biomarkers alone in liquid biopsy, including cfDNA and nipple discharge, remains poorly established. Especially for nipple discharge, which is less widely studied than plasma cfDNA, prior research has focused on cytological or protein markers[23, 24], without systematic characterization of tumor-derived CNV signatures, direct head-to-head comparisons with plasma cfDNA, or rigorous evaluation of its CNV-based diagnostic utility. Most critically, the biological factors governing the release fidelity of tumor-derived CNVs into nipple discharge and their cellular origins remain entirely uncharacterized, hindering the clinical translation of this biofluid.

To address these interconnected gaps, we conducted a systematic analysis of the tumor-derived CNVs in the nipple discharge DNA (ndDNA) in this study. First, we identified and validated 20 recurrent breast cancer CNV features across external datasets, and further verified their ancestral CNV state in a combined scRNA-seq cohort comprising 50 cases across diverse molecular subtypes. We then profiled ndDNA using LP-WGS to assess its diagnostic efficacy and directly compared its performance with that of plasma cfDNA. Finally, we unraveled the cellular and molecular drivers of tumor-derived CNV release at single-cell resolution, with survival analyses linking high CNV fidelity in ndDNA to favorable patient outcomes. Collectively, our findings established nipple discharge as a novel non-blood liquid biopsy platform, provided a clinically actionable CNV-based diagnostic tool, and offered new insights into tumor-biomarker shedding that could inform liquid biopsy development across cancers.

## Methods

### Study Design

We performed a two-stage, cross-platform biomarker study. **Stage 1** mined CNVs from 1,086 primary breast tumors and 942 matched peripheral blood mononuclear cell (PBMC) controls in the TCGA-BRCA cohort to derive a breast-cancer-specific CNV panel. In the subsequent phase, patient-level phylogenetic reconstruction of a multi-subtype single-cell RNA-seq cohort (n=50) confirmed the panel’s features as early oncogenic events conserved across molecular subtypes. **Stage 2** prospectively validated the panel in two independent clinical series: a cohort of 57 women with nipple discharge samples and an independent multicenter set of 183 plasma cfDNA samples. All participants were diagnosed through standard clinical and histopathological evaluation. Breast cancer cases were further classified into clinical stages according to the 8th edition of the American Joint Committee on Cancer (AJCC) staging system[62] and into molecular subtypes based on hormone receptor (HR) and human epidermal growth factor receptor 2 (HER2) status determined by immunohistochemistry. In the first clinical series, all specimens underwent LP-WGS. Single-cell RNA sequencing (scRNA-seq) was then used to analyze matched tumor tissue from five malignant cases to trace the cellular origin of nipple-discharge-derived CNVs.

### Recurrent CNV identification

We downloaded copy-number profiles for 1,086 primary breast cancer tissues and 942 matched PBMC controls from the TCGA-BRCA cohort[63] via the National Cancer Institute Genomic Data Commons portal[64]. Recurrent amplifications and deletions were identified using Genomic Identification of Significant Targets in Cancer 2.0 (GISTIC 2.0, v2.0.23)[25] under default parameter settings (confidence level=0.95, q-value threshold=0.1, amplification threshold=0.1, deletion threshold=-0.1, and arm-level peeling enabled). Output gene-level tables and region summaries were retained for downstream filtering.

### Determination of CNV frequency cutoff

To balance sensitivity and panel size for clinical cost-effectiveness, we modeled the relationship between CNV prevalence and diagnostic sensitivity in the TCGA-BRCA cohort. Starting with a 10% prevalence threshold, we increased the cutoff in 5% increments, recorded the number of retained loci, and computed the corresponding breast cancer detection sensitivity in the same cohort. A 50% prevalence threshold yielded a compact feature set (20 loci) while maintaining ≥98% diagnostic sensitivity, and was therefore selected as the final cutoff.

### Patients

#### The nipple discharge cohort

A prospective cohort of 57 women with nipple discharge (aged 34–70 years) was enrolled at the Yantai Yuhuangding Hospital in Shandong, China (Supplementary Table 1). Nipple discharge samples were collected before treatment. The final pathologic diagnosis identified 39 patients with breast malignancy and 18 patients with benign breast diseases. In a subset of 5 patients with pathologically confirmed malignancy, matched primary tumor tissue and peripheral blood samples were also obtained, enabling comparative multi-omics analysis. The primary tumor tissue samples underwent scRNA-seq, and the extracted cfDNA underwent whole-genome sequencing (WGS).

#### The patient-derived organoid (PDO) cohort

Primary breast cancer biopsies were obtained from the Department of Breast Surgical Oncology, Chinese Academy of Medical Sciences Cancer Hospital. Two patients had HR-positive, HER2-negative breast cancer. Two patients had HR-negative, HER2-positive breast cancer. Two patients had triple-negative breast cancer (TNBC). Each patient had a parallel patient-derived organoid and a patient-derived tissue. Tumor epithelial cells were isolated and cultured as organoids as described by Sato et al.[65]. All 12 samples underwent scRNA-seq.

#### The plasma cfDNA cohort

A multicenter retrospective cohort study was conducted to evaluate the diagnostic performance of plasma cfDNA for breast cancer detection[66]. It comprised 183 female patients (aged 24–80 years), including 149 with breast cancer and 34 with benign breast diseases, recruited from the Cancer Hospital, Chinese Academy of Medical Sciences, and the Yantai Yuhuangding Hospital (Supplementary Table 2). A total of 183 peripheral blood samples were collected before treatment. All samples underwent WGS.

### External datasets for diagnostic performance validation

Three breast cancer datasets (the GSE87048 cohort[26], n = 100, early stage breast cancer; the GSE118527 cohort[27], n = 401; the Molecular Taxonomy of Breast Cancer International Consortium [METABRIC] cohort[28], n = 2,087) and one pan-cancer dataset (the MSK Clinicogenomic Harmonized Oncologic Real-World Dataset [MSK-CHORD] pan-cancer cohort[29], breast cancer [n=1,278], colorectal cancer [n=1,266], prostate cancer [n=715], non-small cell lung cancer [n=1,870], pancreas cancer [n=622]) were enrolled. Processed CNV data were directly obtained from the respective datasets to ensure consistency with the original study protocols. All datasets were normalized to maintain analytical comparability across platforms. A positive diagnosis was defined by the detection of any one of the 20 predefined CNV features, using established calling thresholds and analytical parameters consistent with the original study methodologies. Diagnostic performance was evaluated using sensitivity and specificity, with 95% confidence intervals calculated via Wilson’s methods.

### *The combined cohort* for breast cancer ancestral CNV state validation

A combined cohort comprised two external and two in-house datasets, including Luminal/HR+ (n = 27), HR−/HER2+ (n = 11), and TNBC (n = 12) samples (Supplementary Table 3). Specifically, the external data included two published breast cancer scRNA-seq datasets: GSE161529 (Luminal/HR+ [n = 19], HR−/HER2+ [n = 6], TNBC [n = 8])[67, 68] and GSE199219 (Luminal/HR+ [n = 2], HR−/HER2+ [n = 1], TNBC [n = 2])[69]. The in□house data were derived from *the nipple□discharge cohort* (Luminal/HR + [n = 4], HR−/HER2+ [n = 2]) and *the PDO cohort* (Luminal/HR + [n = 2], HR−/HER2+ [n = 2], TNBC [n = 2]). The gene□expression matrices of the external datasets were downloaded from the Gene Expression Omnibus (GEO) database. All datasets were then integrated and batch□corrected using the Harmony package (v1.2.3)[70] to enable reliable cross□cohort analysis.

### Single-cell RNA sequencing

#### Library construction and sequencing

Fresh primary breast cancer tissues were surgically harvested and immediately washed in ice-cold PBS (SH30256.01, Hyclone, Logan, UT, USA), then dissociated using the SeekMate Tissue Dissociation Reagent Kit A Pro (K01801-30, SeekGene, Beijing, China) according to the manufacturer’s instructions. DNase Ⅰ (9003-98-9, Sigma-Aldrich, St. Louis, MO, USA) treatment was applied as needed based on homogenate viscosity. After erythrocyte removal (R1010, Solarbio, Beijing, China), cell count and viability were assessed using a Fluorescence Cell Analyzer (Rigel S2, Countstar, Shanghai, China) with AO/PI reagent, followed by optional debris and dead cell removal (130-109-398/130-090-101, Miltenyi, Bergisch Gladbach, NRW, Germany). Fresh cells were washed twice in RPMI1640 (11875119, Gibco, Waltham, MA, USA) and resuspended at 1×10□cells/ml in RPMI1640 with 2% FBS (10100147C, Gibco, Waltham, MA, USA). Single-cell RNA-seq libraries were prepared using the SeekOne® DD Single Cell 3’ library preparation kit (K00202, SeekGene, Beijing, China). Cells were mixed with reverse transcription reagent and loaded into the SeekOne® chip S3, after which Barcoded Hydrogel Beads and partitioning oil were added. After emulsion droplet generation, reverse transcription was performed at 42°C for 90 minutes, followed by inactivation at 85°C for 5 minutes. cDNA was purified, amplified, cleaned, fragmented, end-repaired, A-tailed, and ligated to sequencing adaptors. Indexed polymerase chain reaction amplified the 3’ polyA regions containing Cell Barcodes and Unique Molecular Indices. Libraries were cleaned with VAHTS DNA Clean Beads (N411-01, Vazyme Biotech, Nanjing, JS, China), quantified using Qubit (Q33226, Thermo Fisher Scientific, Waltham, MA, USA) and Bio-Fragment Analyzer (Qsep400, Bioptic, Hsinchu, TW, China), and sequenced on Illumina NovaSeq 6000 with PE150 read length. The scRNA-seq reads from 5 samples were aligned to the human reference genome GRCh38 using the Cell Ranger (v7.1.0)[71].

#### Data processing, batch correction, and clustering

Subsequent single-cell data analysis was conducted using the Seurat package (v4.4.0)[72]. Quality control filtering was performed with the following parameters: cells expressing between 100 and 7,000 genes were retained, and those with mitochondrial gene content exceeding 10% were excluded. Following filtering, the gene expression matrices were normalized and scaled using Seurat’s “*NormalizeData*” and “*ScaleData*” functions. To mitigate batch effects across patient samples, the Harmony package (v1.2.3) was used for data integration[70]. A total of 30,709 genes were retained for downstream analysis, encompassing 48,996 cells from primary breast cancer samples. Principal component analysis (PCA) was conducted on the batch-corrected expression matrix, using highly variable genes identified by the Seurat “*FindVariableGenes*” algorithm. Dimensionality reduction was performed via the “*RunPCA*” function. Cell clusters were subsequently identified by applying the “*FindClusters*” function to the most informative principal components.

#### Cell type annotation by canonical markers

The marker genes for each cell type were listed as follows: CD3D and CD3E for T cells (CD4, CD8A, and FOXP3 for further grouping); JCHAIN, MZB1, and IGKC for plasma cells; NKG7, GNLY, and NCAM1 for natural killer cells; LYZ, CD14, and CD68 for myeloid cells; TPSAB1, TPSB2, and CPA3 for mast cells; EPCAM, CDH1, and KRT8 for epithelial cells; PECAM1, VWF, and CD34 for endothelial cells; HLA-DRA and CD83 for dendritic cells; PDGFRA, PDGFRB, and COL1A1 for fibroblasts; MS4A1 and CD79A for B cells[73, 74].

#### CNV inference

The inferCNV package (v1.19.1)[30] was used to infer large-scale CNVs in epithelial subclusters. Downsampled T cells and myeloid cells served as reference populations to normalize gene expression and identify CNV events. The analysis was performed within a single patient using the hidden Markov model. The “*subcluster*” mode was used to determine clonal CNV changes.

### Phylogenetic tree construction and ancestral CNV state inference

Phylogenetic trees for individual patients were reconstructed from the inferCNV data using maximum parsimony implemented in the phangorn package (v2.12.1)[36] in R. Ancestral CNV states for each gene were then inferred using the ancestral.pars function from the phangorn package with the maximum parsimony reconstruction (MPR) algorithm. The algorithm identified the most parsimonious state assignments and computed their associated likelihoods at every internal node. An ancestral state was considered confidently resolved when its maximum parsimony likelihood exceeded 0.5, the conventional threshold for binary classification in parsimony-based reconstruction.

### Calculation of Cohen’s d effect sizes

To quantify the functional differences associated with each CNV feature, all epithelial cells from *the combined cohort* were stratified into two groups based on whether the CNV feature was present or absent. Gene Ontology Biological Process (GOBP) pathway scores were calculated for each cell using the Seurat AddModuleScore function with default parameters. Cohen’s d effect sizes were then computed for each GOBP pathway using the effsize package (v0.8.1)[37] in R, with positive values indicating higher pathway activity in CNV feature-positive cells and negative values indicating lower activity. Statistical significance was assessed using the Wilcoxon rank-sum test with Benjamini-Hochberg correction for multiple comparisons.

### Nipple discharge sample processing

Nipple discharge samples were manually extruded, and approximately 0.3–0.5 mL of fluid was collected using a 1-mL syringe. The collected fluid was immediately transferred to 3 mL of preservation solution and stored at 0–4 °C. All samples were processed for library construction and sequencing within 48 hours of collection.

The ndDNA was extracted using the Amp Genomic DNA Kit (TIANGEN Biotech, Beijing, China) following the manufacturer’s instructions. Sequencing libraries were prepared using the NEBNext Ultra II DNA Library Prep Kit (New England Biolabs, Ipswich, MA, USA) with a mean input of 251.9 ng genomic DNA per sample, followed by LP-WGS. The DNA fragments were ligated with 8□bp barcoded sequencing adapters and amplified by PCR. Purified libraries were subjected to massively parallel sequencing on an Illumina HiSeq X Ten platform (Illumina, San Diego, CA, USA). Approximately 4□Gb of raw sequencing data per sample were filtered and aligned to the human reference genome (GRCh37) using BWA-MEM (v0.7.17)[75]. Genomic coverage was quantified with SAMtools mpileup (v1.15)[76]. Coverage values for each 200-Kb genomic bin were calculated and normalized for GC content. An external normal control dataset from healthy individuals was used as a reference baseline for subsequent normalization. A Z-score approach was applied to identify regional CNVs. The thresholds (Z-score > 0.4 for amplification and < −0.8 for deletion) were established based on our preliminary data analysis.

### Plasma cfDNA sample processing

A total of 10 mL of peripheral blood was collected from 188 patients and placed in Streck cfDNA blood collection tubes (Streck, Omaha, NE, USA). Of these, 5 were from the nipple discharge cohort and 183 from *the plasma cfDNA cohort*. Plasma separation was performed using a two-step centrifugation protocol: initial centrifugation at 1,600 × g for 15 minutes at 4°C to isolate plasma from cellular components, followed by a second centrifugation at 16,000 × g for 10 minutes at 4°C to remove residual platelets and debris. The purified plasma was carefully aliquoted and stored at −80°C until analysis.

cfDNA was extracted from plasma samples using the QIAamp Circulating Nucleic Acid Kit (Qiagen, Hilden, Germany) according to the manufacturer’s specifications. The integrity and concentration of the extracted cfDNA were assessed. Library preparation was performed using the Fast Library Prep Kit v2.0 (iGeneTech, Beijing, China) for end repair and adapter ligation. A total of 750 ng of the pre-library was subjected to hybridization capture with the TargetSeq® Pan-Cancer Panel and the TargetSeq One® Hyb & Wash Kit v2.0 (iGeneTech, Beijing, China). The captured libraries were sequenced on the HiSeq X Ten platform (Illumina, San Diego, CA, USA) using PE150. Raw sequencing data were processed with Fastp for quality control and filtering. Clean reads were aligned to the human reference genome (GRCh37) using BWA-MEM (v0.7.17) [75], and CNV calling was performed using CNVnator (v0.4.1)[77]. The thresholds (segment copy-number altered > 0.2 for amplification and < −0.2 for deletion) were established based on our preliminary data analysis.

### Reference genome conversion

To enable comprehensive multi-omics and multi-database analysis, this study converted genomic coordinates between GRCh37 and GRCh38 using the “liftOver” function from the rtracklayer package (version 1.62.0).

### CNV profile visualization and significance testing

CNV profiles per sample were visualized using the cnSpec function from the GenVisR package (v1.34.0)[78]. The alternation frequencies of *the nipple discharge cohort* and *the plasma cfDNA cohort* were displayed using the cnFreq function. The alternative significance of these two cohorts was assessed using the two-proportion Z-test, adjusted for multiple comparisons using the Benjamini-Hochberg (BH) Correction, and visualized with the ggplot2 package (v3.5.1)[79].

### Functional annotation of epithelial subclusters

Epithelial cells were unsupervised re-clustered and combined with GOBP functional enrichment scoring[80] to identify functionally distinct subclusters. The analysis was conducted using the “*AddModuleScore*” function from the Seurat package, which calculates pathway activity scores for each cell based on the average expression of genes within predefined GOBP gene sets. We focused on biological processes relevant to epithelial cell function, including cellular development, epithelial-mesenchymal transition, adhesion, cellular response, OP, and immune response pathways. Pathway scores were computed for each epithelial subcluster, and significantly enriched GOBP terms (adjusted *p*-value < 0.05) were identified with BH correction for multiple comparisons. The resulting pathway activity profiles were visualized using heatmaps to illustrate the functional heterogeneity among epithelial subclusters.

### Fidelity Rate and Fidelity Score calculation

The *Fidelity Rate* is defined as the proportion of predefined CNV features that are jointly detected in both the ndDNA and the subcluster’s inferCNV profile, relative to the total number of features identified in either source.

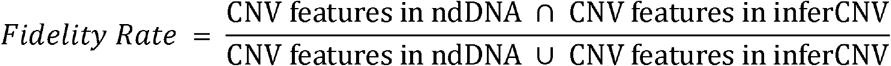

The *Fidelity Score* was derived from a panel of 30 genes identified as the most differentially expressed between groups with high and low Fidelity Rate, reflecting tumor cell shedding propensity. This score was designed to be extensible, enabling its application to external datasets to robustly quantify the underlying biological phenotype across diverse sample populations.

### Survival Analysis

Survival analysis was conducted using the survival (v3.8-3) and survminer (v0.5.0) packages in R. Overall survival (OS) was defined as the time from diagnosis to death from any cause. Disease-free survival (DFS) was defined as the time from diagnosis to disease recurrence or death from any cause. Patients who were alive or lost to follow-up were censored at the last follow-up date. Kaplan-Meier (K-M) survival curves were generated using the survfit function from the survival package, with the formula specified as Surv(time, status) ∼ group_variable, where time denotes survival time and status indicates event status (1 = event, 0 = censored). Survival curves were visualized using the ggsurvplot function from the survminer package. Log-rank tests were performed using the survdiff function to compare survival distributions between groups. All statistical tests were two-sided, and *p*-values < 0.05 were considered statistically significant.

### Statistical analysis

Statistical analyses were performed using R software (v4.4.3). Between-group comparisons were made using two-tailed Student’s t-tests (for two groups) or one-way/two-way ANOVA (for three or more groups). For non-normally distributed data, the Wilcoxon signed-rank test was employed. Correlations were analyzed using Spearman’s method. Statistical significance was defined as a two-sided *p*-value < 0.05. The BH method was used to calculate adjusted *p*-values for multiple comparisons. The receiver operating characteristic (ROC) curves for the CNV features diagnostic model in *the nipple discharge cohort* and *the plasma cfDNA cohort* were generated using the pROC package in R.

## Results

### Identification of recurrent CNV features for breast cancer

To identify recurrent somatic CNV features in breast cancer, we analyzed whole-genome CNV data from 1,086 primary breast tumors and 942 matched PBMC controls in the TCGA-BRCA cohort. Using the GISTIC 2.0 analytical pipeline[25], we identified 158 recurrent somatic CNV features (87 amplifications and 71 deletions) specifically associated with breast carcinogenesis (Supplementary Fig. S1A). We then applied a multi-step filtering strategy to derive a concise feature panel. First, 135 features with an adjusted *p*-value < 0.01 were retained (75 amplifications and 60 deletions). Next, we applied a population frequency threshold of 50% to optimize the balance between panel size and diagnostic sensitivity (Supplementary Fig. S1B and S1C), yielding 22 features (14 amplifications and 8 deletions). Finally, to ensure biological relevance, we excluded two features that did not encompass known protein-coding genes or cancer-related elements, yielding a final panel of 20 high-confidence CNV features (12 amplifications and 8 deletions) for all subsequent analyses (**Fig. 1A**; Supplementary Table S4). CNV features exhibited clustered genomic distributions, with amplifications enriched on chromosomes 1q (6 features) and 8q (5 features), and deletions clustered on chromosomes 8p, 16q, and 17p (**Fig. 1B**).

**Figure 1.**
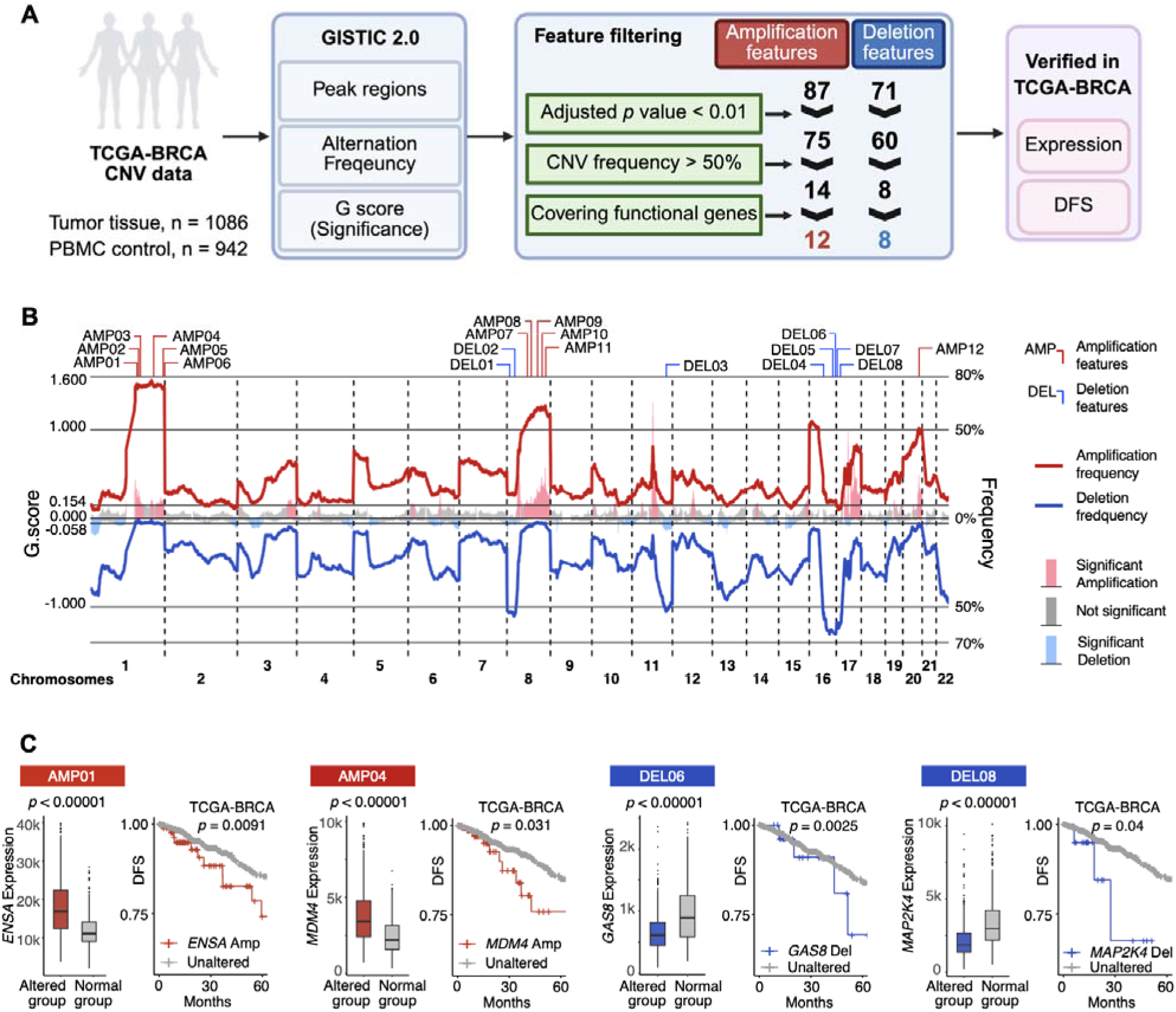
Identification and validation of copy-number variation (CNV) features in breast cancer. **A,** Workflow for CNV feature selection. Genomic Identification of Significant Target in Cancer 2.0 (GISTIC 2.0, v2.0.23) was performed on The Cancer Genome Atlas Breast Invasive Carcinoma (TCGA-BRCA; n=1086), identifying 158 recurrent CNV features. These were filtered to 12 amplifications and 8 deletions for subsequent functional validation within th same cohort. PBMC, peripheral blood mononuclear cell; DFS, disease-free survival. **B,** CNV landscape of the TCGA-BRCA cohort. The line plot shows the frequency of CNVs across the genome. Genomic regions with significant alterations are highlighted as colored bars. The locations of the 20 selected CNV features are annotated above the plot. **C,** Functional validation of CNV features. Observed CNVs were concordant with changes in mRNA expression of the functional genes they covered. Patients with specific functional single-gene CNVs exhibited significantly worse DFS.

To validate their biological and clinical relevance, we conducted two lines of investigation. First, integrated genomic analysis confirmed that CNVs in genes within these features were significantly correlated with corresponding mRNA expression levels (i.e., amplifications with overexpression and deletions with underexpression) (**Fig. 1C**; Supplementary Fig. S1D). Second, survival analysis showed that patients with amplifications of key oncogenes or deletions of tumor suppressor genes encompassed by selected CNV features had significantly worse DFS (**Fig. 1C**).

### Validation of the 20-CNV feature panel’s diagnostic performance in external cohorts

Before assessing the utility of the 20 CNV features in nipple discharge, we validated them as breast cancer-specific biomarkers in multiple independent tissue cohorts. In the GSE87048 cohort[26] (n = 100, early stage), the CNV features detected breast cancer with 77.0% sensitivity (95% CI: 67.9%–84.2%) and minimal background signal in matched PBMCs (1/100 positive), confirming their cancer-specific origin (**Fig. 2A**). The high sensitivity was further confirmed in the larger GSE118527 cohort[27] (92.8%, 95% CI: 89.8%–94.9%; n = 401) and the METABRIC cohort[28] (93.7%, 95% CI: 92.6%-94.7%; n = 2,087) (**Fig. 2A and B**).

**Figure 2.**
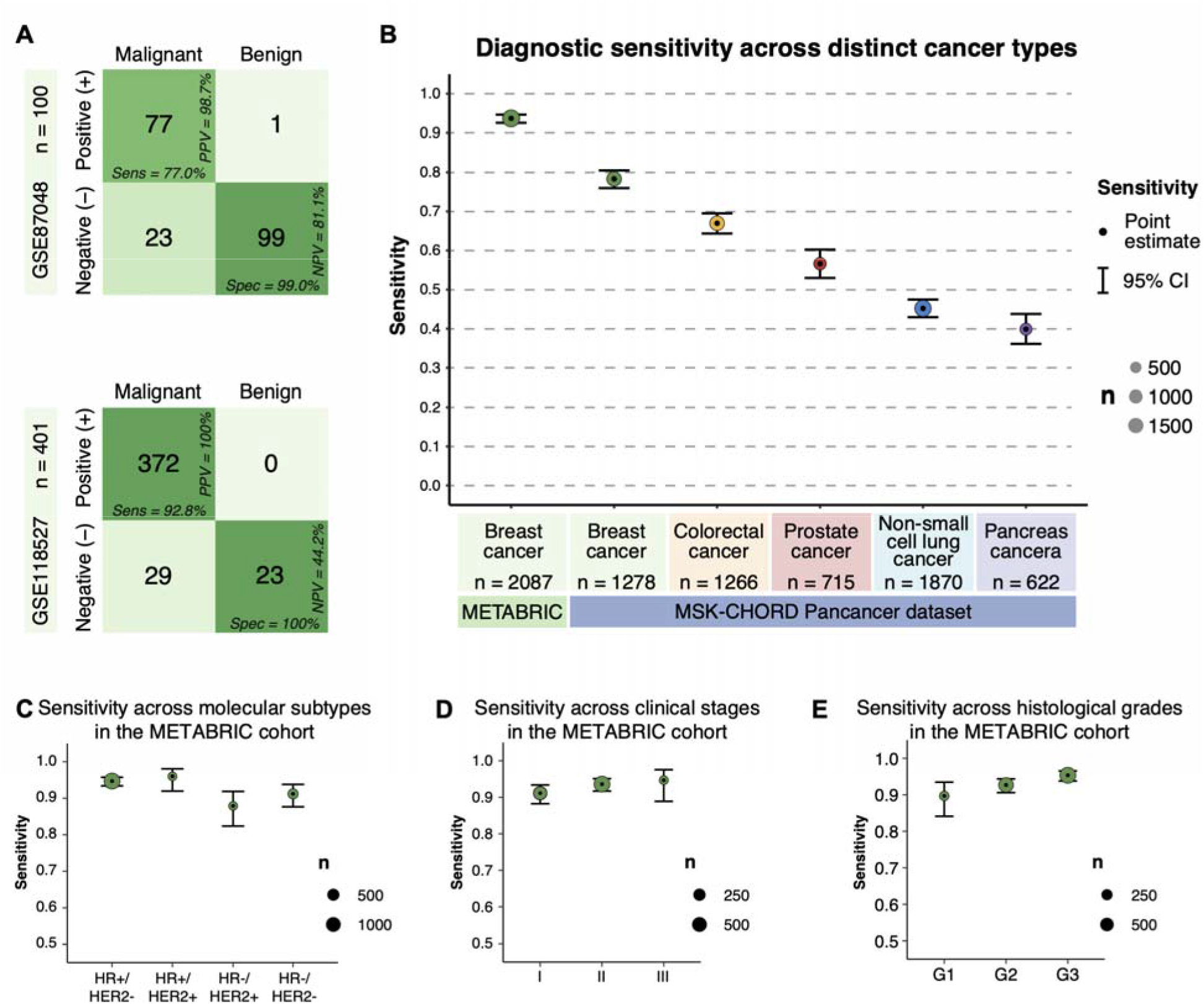
External validation of the CNV features diagnostic model. **A,** Confusion matrices for the GSE87048 cohort (100 early-stage breast cancer patients with paired cancer tissue and PBMC normal controls) and the GSE118527 cohort (401 breast cancer patients with cancer tissue, 23 of whom had PBMC normal controls). **B,** Sensitivity of the CNV features diagnostic model in the Molecular Taxonomy of Breast Cancer International Consortium (METABRIC) cohort (2087 patients with breast cancer tissue) and the MSK Clinicogenomic Harmonized Oncologic Real-World Dataset (MSK-CHORD) pan-cancer cohort (breast cancer [n=1278], colorectal cancer [n=1266], prostate cancer [n=715], non-small cell lung cancer [n=1870], pancreas cancer [n=622]), grouped by cancer type. The point estimates (dots) are shown with 95% confidence intervals (error bars). The size of each dot corresponds to the number of patients in the subgroup. **C-E,** Sensitivity of the CNV features diagnostic model across patient subgroups in the METABRIC cohort, stratified by molecular subtype (**C**), clinical stage (**D**), and histological grade (**E**). The point estimates (dots) are shown with 95% confidence intervals (error bars). The size of each dot corresponds to the number of patients in the subgroup.

To evaluate the cancer-type specificity of these features, we performed a pan-cancer analysis using the MSK-CHORD dataset[29]. The features showed the highest sensitivity in breast cancer (78.3%, 95% CI: 0.760–0.805) compared with other major cancer types (e.g., colorectal cancer: 67.0%; prostate cancer: 56.6%), supporting their relative specificity for breast malignancies (**Fig. 2B**). We further investigated the impact of clinical parameters on detection sensitivity using the METABRIC cohort. The CNV features maintained robust performance across all molecular subtypes, with the lowest sensitivity still observed at 87.9% in the HR-negative/HER2-positive subgroup (**Fig. 2C**). Sensitivity remained consistently high across clinical stages and histological grades, with a positive trend toward more advanced disease, supporting the biological relevance of these CNV features in tumor progression (**Fig. 2D and E**). This robust sensitivity was also maintained across variations in patient age, tumor size, and histological type (Supplementary Fig. S2A). Collectively, these analyses establish our 20 CNV features as highly sensitive and specific biomarkers for breast tumor tissue, providing a solid foundation for their subsequent application in liquid biopsy.

### Recurrent and ancestral CNV features drive conserved functional programs across breast cancer subtypes

To elucidate the evolution of copy-number and its functional consequences across breast cancer molecular subtypes, we integrated single-cell inferCNV and transcriptome analyses in the combined cohort (n = 50), encompassing Luminal/HR+ (n = 27, 26 HR+/HER2− and one HR+/HER2+), HR−/HER2+ (n = 11), and TNBC (n = 12) subtypes (**Fig. 3A**; Supplementary Fig. S3A). Cell types were annotated based on marker gene expression (Supplementary Fig. S3B). Epithelial cells from different molecular subtypes showed distinct distribution patterns in Uniform Manifold Approximation and Projection (UMAP) plots (**Fig. 3B**).

**Figure 3.**
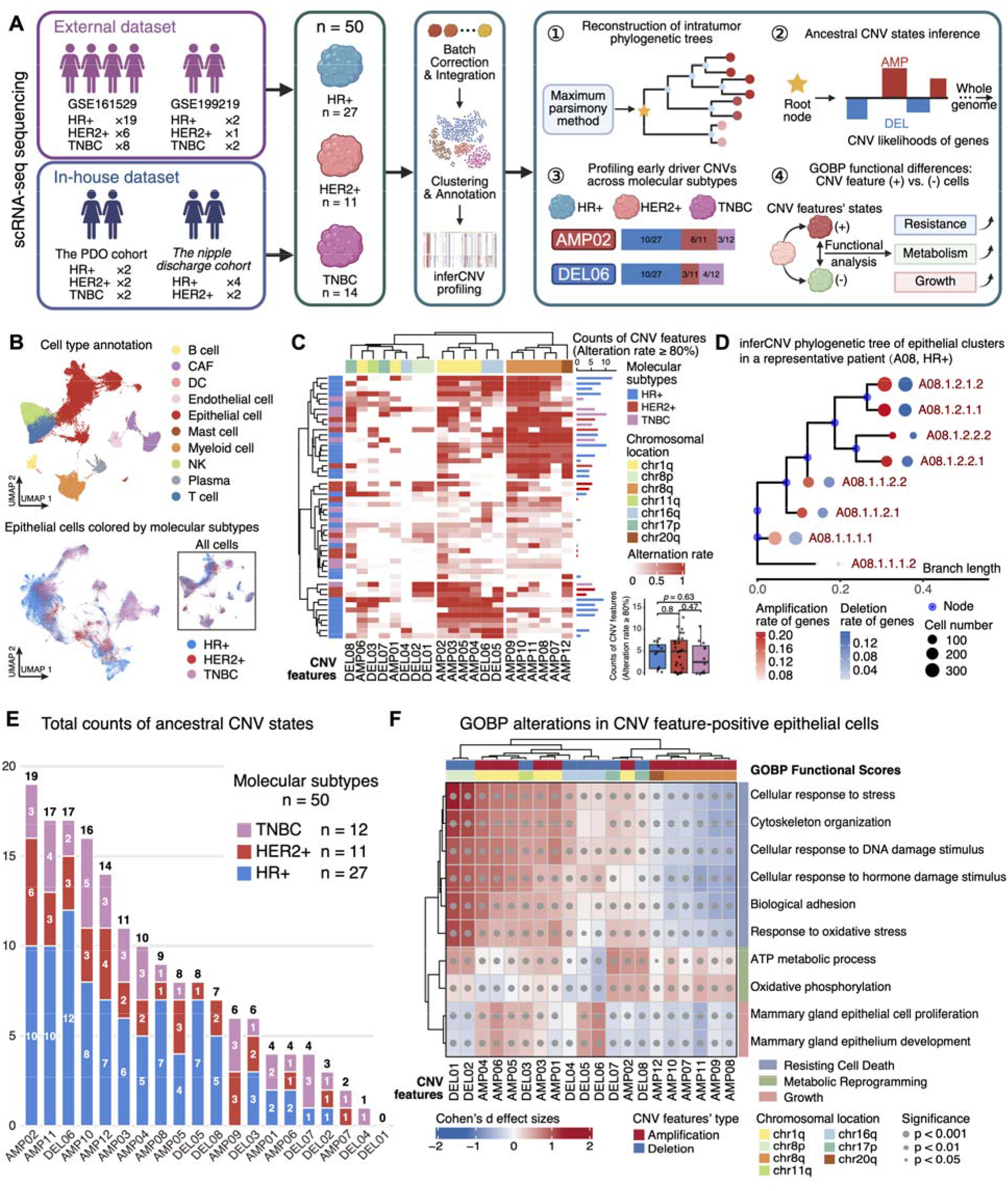
Integrative single-cell inferCNV profiling across molecular subtypes identifies early driver CNVs and their functional consequences. **A,** Study design and analytical workflow. **A** was generated using cBioPortal (https://www.cbioportal.org/). **B,** Uniform Manifold Approximation and Projection (UMAP) visualization of integrative single-cell transcriptomes, colored by cell type and molecular subtype. **C,** Heatmap showing the fraction of epithelial cells carrying each CNV feature across 50 patients, annotated by molecular subtype, with an accompanying box plot showing counts of dominant CNV features across these subtypes. **D,** InferCNV-based phylogenetic tree of epithelial cell clusters from a representative patient (A08, HR+). **E,** Total counts of ancestral CNV states across 20 CNV features, stratified by molecular subtype in the combined cohort. **F,** Heatmap of Cohen’s d effect sizes quantifying Gene Ontology Biological Process (GOBP) pathway activity differences between CNV feature-positive and −negative epithelial cells across 20 CNV features, annotated by significance level and chromosomal location.

Next, we performed inferCNV analysis[30] on epithelial cells from individual patients across the three molecular subtypes to delineate subclonal architectures (**Fig. 3A**). For each patient, we calculated the proportion of epithelial cells carrying each of the 20 CNV features and then performed unsupervised clustering. This analysis revealed that 35 of 50 patients harbored at least one high-alteration CNV feature (alteration rate >80%). No significant differences were observed in the counts of high-alteration CNV features among the three subtypes. Certain CNV features showed notable co-occurrence, most prominently AMP07∼12, which localize to chromosomes 8q and 20q (**Fig. 3C**). This co-occurrence pattern has also been reported in previous studies[31–33], and its biological effect may stem from a functional cooperative interaction between MYC and ZNF217[34, 35].

To validate whether the 20 CNV features represent early oncogenic drivers in breast cancer, we reconstructed phylogenetic trees from inferCNV data for individual patients and inferred ancestral CNV states[36]. A consistent trend across all 50 patients’ phylogenetic trees was that more evolutionarily distant cell populations had higher rates of gene alterations and larger cell counts (**Fig. 3D**; Supplementary File S1). There were no obvious differences in ancestral CNV state patterns among the three subtypes (Supplementary Fig. S3C). Most CNV features (19/20) were detected in the ancestral CNV state, with four CNV features present in > 30% of the combined cohort (**Fig. 3E**). The 20 CNV features exhibited distinct distribution patterns within epithelial cells (Supplementary Fig. S3D). We assessed their functional impact by comparing enriched GOBP terms between epithelial cells with and without each CNV feature, using Cohen’s d effect sizes[37]. Unsupervised clustering of these functional profiles delineated three predominant oncogenic modules: cell death resistance, metabolic reprogramming, and mammary gland□specific growth signaling. The co occurring CNV cluster AMP07∼12, previously identified on chromosomes 8q and 20q in our above analysis, was functionally linked to a significant upregulation of ATP metabolic processes and oxidative phosphorylation (**Fig. 3F**), underscoring its role in promoting tumor bioenergetic capacity.

### Diagnostic performance of CNV features in nipple discharge outperforms plasma cfDNA

We next evaluated the diagnostic performance of the 20 CNV features in the intended clinical samples: nipple discharge and plasma cfDNA. In these two cohorts, we applied LP-WGS to nipple discharge samples from 57 patients (39 malignant, 18 benign) and WGS to plasma cfDNA samples from 183 patients (149 malignant, 34 benign) (**Fig. 4A**).

**Figure 4.**
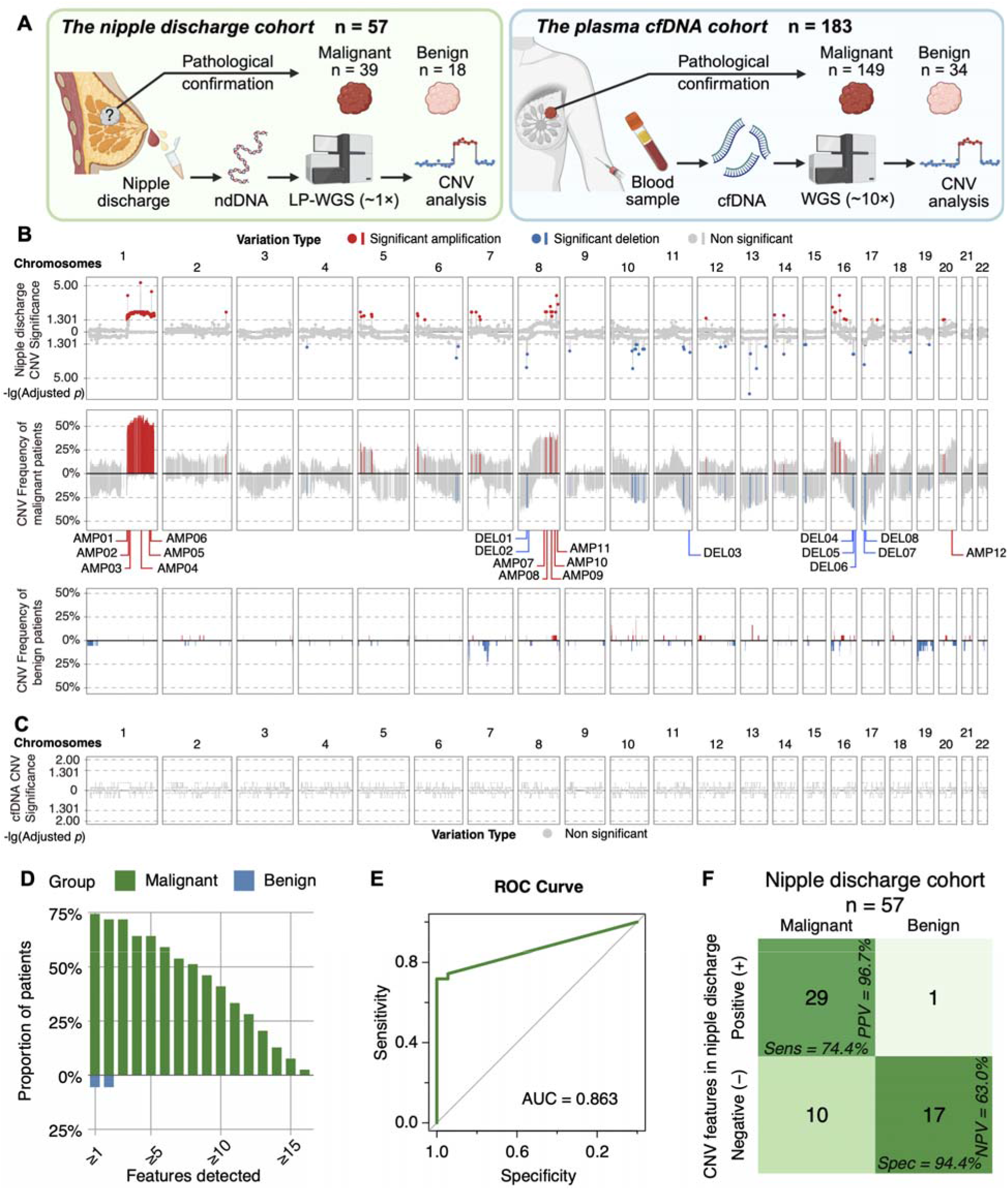
Performance of the CNV features diagnostic model in *the nipple discharge* and *plasma cfDNA cohorts*. **A,** Study design and analytical workflow for both cohorts. **A** was generated using cBioPortal (https://www.cbioportal.org/). ndDNA nipple discharge DNA, LP-WGS low-pass whole-genome sequencing, cfDNA cell-free DNA, WGS whole-genome sequencing. **B,** Genome-wide CNV landscape of the *nipple discharge* cohort. The top panel displays −log10(adjusted *p*-values) for differences in CNV frequency between malignant and benign patients; significant regions are colored red or blue. The middle and bottom panels display CNV frequencies in malignant and benign patients, respectively. Annotations mark the genomic positions of the 20 CNV features. **C,** −log10(adjusted *p*-values) for CNV-frequency differences in *the plasma cfDNA cohort*; no region reached statistical significance. **D,** Proportion of positive calls in malignant and benign patients in *the nipple discharge cohort* at increasing detection-threshold numbers of CNV features. **E,** Receiver operator characteristic (ROC) curve for *the nipple discharge cohort*; area under the ROC curve (AUC) = 0.863 (95 % CI 0.788–0.938). **F,** Confusion matrices illustrating the diagnostic performance of the CNV features diagnostic model in *the nipple discharge cohort* (39 malignant and 18 benign patients).

Analysis of nipple discharge revealed distinct CNV landscapes between malignant and benign cases. Malignant samples exhibited a significantly higher burden of genome-wide CNVs (Supplementary Fig. S4A and S4B). Critically, the CNV profiles closely recapitulated the selected CNV features, with significant amplifications on chromosomes 1q and 8q (encompassing 11 of the 12 amplification features) and deletions covering all 8 deletion features (**Fig. 4B**). In contrast, CNV analysis of plasma cfDNA showed no significant differences between malignant and benign groups **(Fig. 4C)**, highlighting that nipple discharge is a superior biospecimen for CNV detection.

We then quantified the diagnostic accuracy of the CNV features in *the nipple discharge cohort*. ROC analysis of the model, which employed a simple positive-calling criterion (positivity defined as the detection of any CNV feature), yielded an AUC of 0.863 (95% CI: 0.788–0.938), indicating strong discriminative power (**Fig. 4D and E**). Under this criterion, the model attained a sensitivity of 74.4% (95% CI: 58.9%–85.4%), a specificity of 94.4% (95% CI: 74.2%–99.7%), a positive predictive value (PPV) of 96.7%, and a negative predictive value (NPV) of 63.0% (**Fig. 4F**). Moreover, the model maintained high sensitivity across patient subgroups in the nipple discharge cohort (stratified by molecular subtype, clinical stage, and histological grade), demonstrating its broad applicability across diverse patient types (Supplementary Fig. S4C). For *the plasma cfDNA cohort*, the ROC analysis yielded an AUC of 0.517 (95% CI: 0.474–0.560), with a sensitivity of 100.00% and a specificity of 5.88% (Supplementary Fig. S4D). Delong’s test confirmed that the AUC of *the nipple discharge cohort* was significantly higher than that of *the plasma cfDNA cohort* (*p* < 0.0001), confirming the superior diagnostic performance of nipple discharge. With high specificity and PPV, this assay serves as an excellent rule-in test, reliably confirming disease when positive. In contrast, its satisfactory sensitivity suggests it could fit into a broader, multi-model diagnostic strategy without excessive false negatives.

### Single-cell analysis of CNV features in nipple discharge reveals cellular origins and release fidelity

To investigate the cellular origins of CNV features in nipple discharge and the factors influencing their detection, we established a multi-modal analytical framework. We prospectively collected matched samples from five breast cancer patients (P61, P62, P63, P64, P66) in *the nipple discharge cohort*, including: (i) primary tumor tissue for scRNA-seq (Supplementary Fig. S5A); (ii) nipple discharge for LP-WGS of the ndDNA; and (iii) peripheral blood for WGS of plasma cfDNA (**Fig. 5A**).

**Figure 5.**
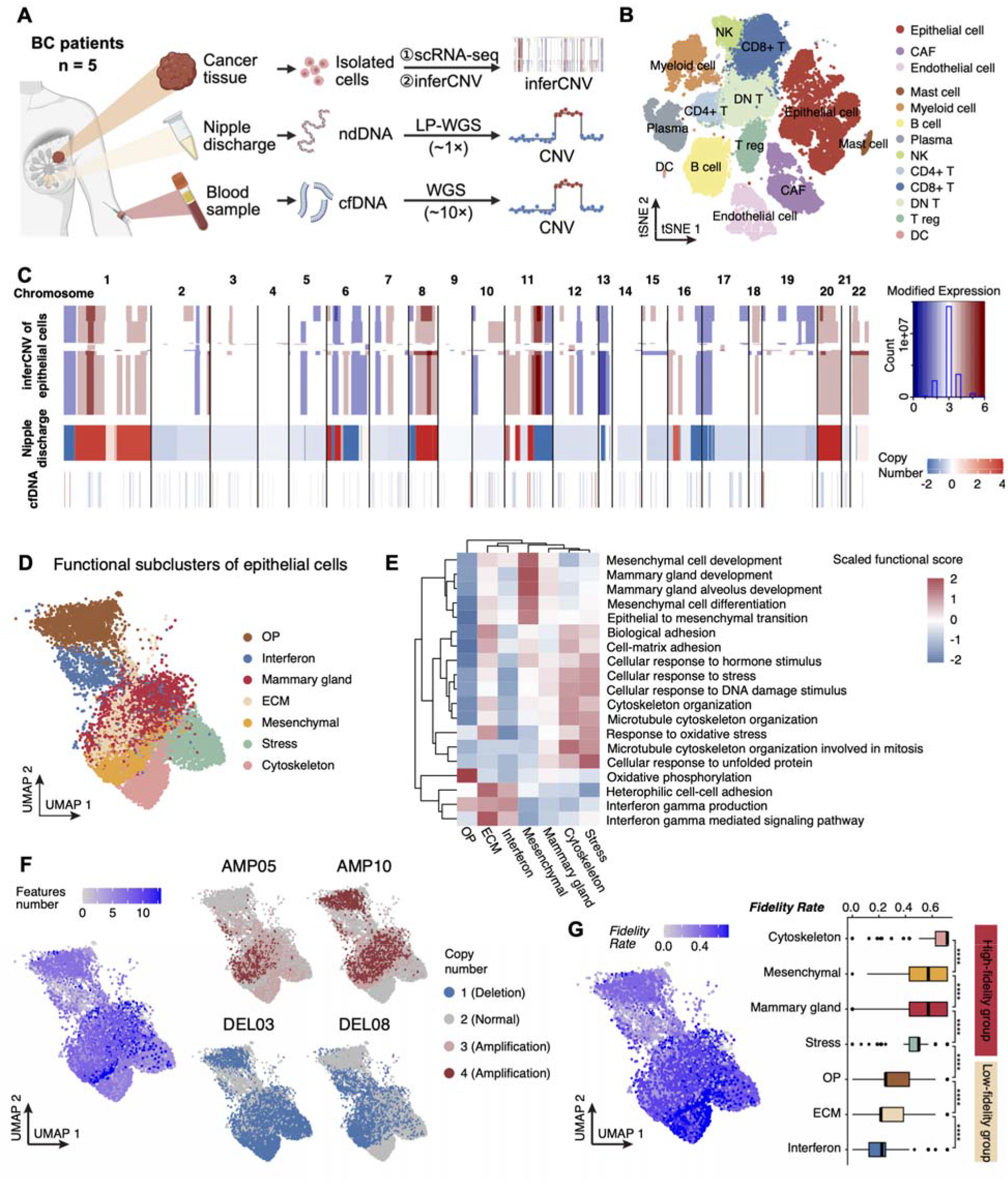
Single-cell tracing of the cellular origins of CNV features in nipple discharge. **A**, Integrated multi-omics study design using matched clinical samples. Schematic overview of paired sample collection (tumor, nipple discharge, blood) and corresponding genomic analyses in five breast cancer patients from the nipple discharge cohort. BC, breast cancer; scRNA-seq, single-cell RNA sequencing. **A** was generated using cBioPortal (https://www.cbioportal.org/). **B,** t-distributed Stochastic Neighbor Embedding (t-SNE) visualization of single-cell transcriptomes, colored by cell type. CAF, cancer-associated fibroblast; NK, natural killer cell; DN T, double-negative T cell; T reg, regulatory T cell; DC, dendritic cell. **C,** Genome-wide heatmaps comparing the inferCNV profile of epithelial subclusters with CNV profiles derived from nipple discharge and cfDNA samples in a representative patient (P62). **D,** UMAP visualization of seven identified functional epithelial cell subclusters based on transcriptomic signatures. OP, oxidative phosphorylation; ECM, extracellular matrix. **E,** Heatmap showing expression levels of key functional gene sets across the seven functional epithelial cell subclusters. **F,** UMAP visualization of epithelial cell subclusters, colored by the number of features detected per cell and the copy-number state of representative CNV features. **G,** UMAP visualization of epithelial cell subclusters, colored by the *Fidelity Rate* calculated per cell. The box plot shows the *Fidelity Rate* across the seven functional epithelial cell subclusters, stratified into high- and low-fidelity groups.

scRNA-seq analysis of tumor tissues first delineated the tumor microenvironment, identifying malignant epithelial cells and diverse stromal and immune populations (**Fig. 5B**; Supplementary Fig. S5B). The inferCNV profiles for malignant epithelial cells were then reconstructed using non-malignant cells as a reference baseline. We assessed the global concordance of CNV profiles across these compartments. The CNV landscape derived from ndDNA showed remarkable similarity to the inferCNV profile reconstructed from scRNA-seq. In contrast, the plasma cfDNA profile showed poor concordance with the tumor baseline, despite deeper sequencing coverage (∼10× vs. ∼1× for nipple discharge), underscoring nipple discharge as a superior liquid biopsy source for capturing tumor-derived genomics (**Fig. 5C**; Supplementary Fig. S5C).

To identify which tumor cells contribute most to the nipple discharge signal, we classified malignant epithelial cells into seven functional subclusters based on transcriptomic profiles: Oxidative phosphorylation (OP), Interferon, Mammary gland, Extracellular matrix (ECM), Mesenchymal, Stress, and Cytoskeleton (**Fig. 5D and E**). Evaluation of the 20 predefined CNV features across epithelial subclusters revealed significant heterogeneity in both the number and types of features. However, each subcluster maintained a high degree of similarity, directly linking specific CNV patterns to distinct cellular phenotypes (**Fig. 5F**; Supplementary Fig. S5D).

To quantify how faithfully each epithelial subcluster delivers its CNV features into nipple discharge, we developed a metric called the *Fidelity Rate*. Based on this metric, the subclusters formed two distinct groups: the Cytoskeleton, Mesenchymal, Mammary Gland, and Stress subclusters formed a high-fidelity group with significantly higher rates, whereas the OP, ECM, and Interferon subclusters formed a low-fidelity group (**Fig. 5G**).

### Functional mechanisms and clinical prognostic value of the epithelial cell *Fidelity Score*

To further establish a molecular metric reflecting tumor cell shedding propensity, we defined the *Fidelity Score* (Supplementary Table S5). It is calculated from the top 30 differentially expressed genes between groups with high or low *Fidelity Rate* (**Fig. 6A**). Our comprehensive characterization of the *Fidelity Score* incorporated three key dimensions: its association with genomic, functional, and spatial states, its relationship with cell–cell interactions, and its clinical relevance.

**Figure 6.**
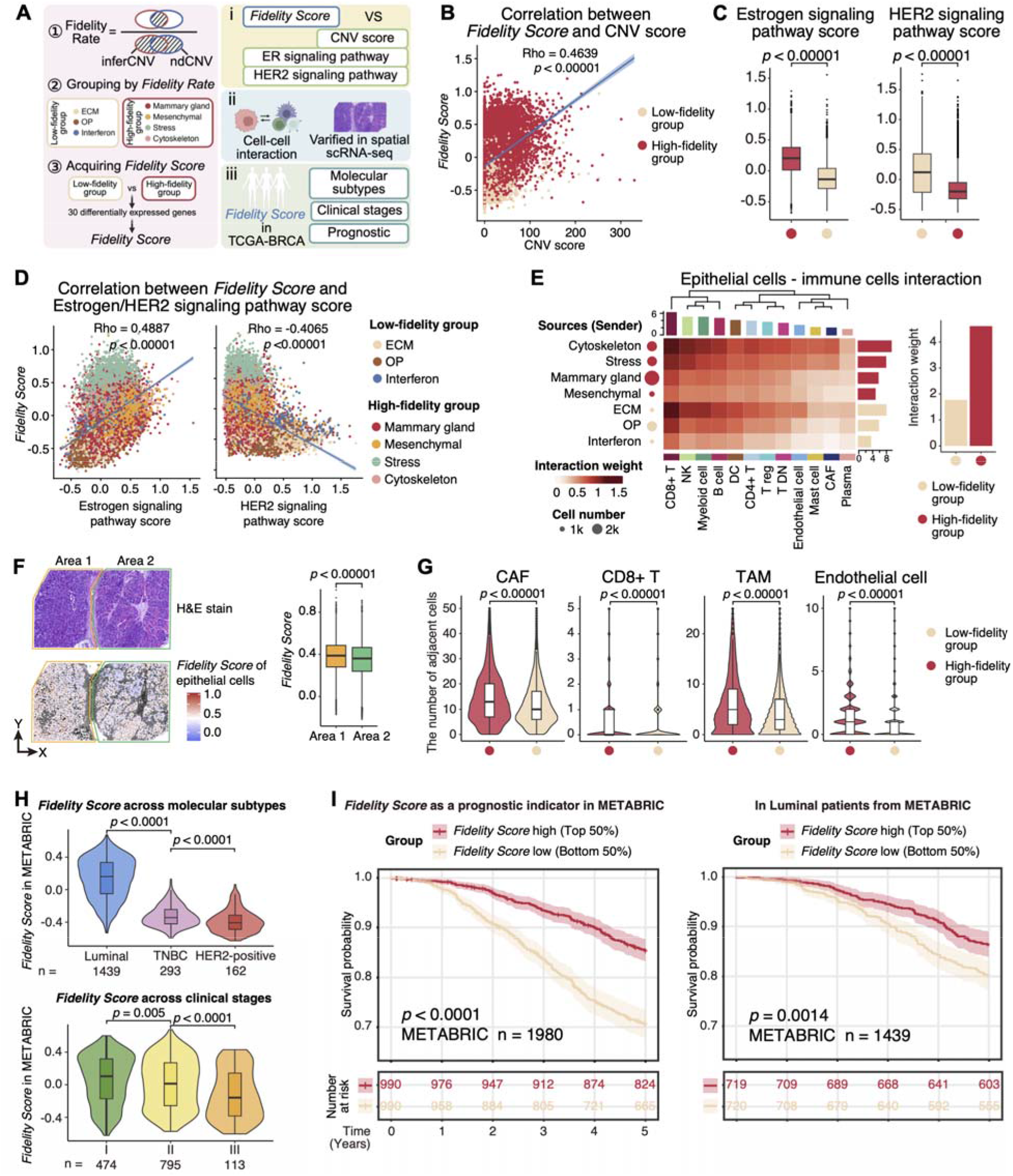
Functional characterization of high-fidelity epithelial cell groups and the *Fidelity Score* as a prognostic indicator in breast cancer. **A**, Workflow for calculating the *Fidelity Score* and its subsequent analysis to elucidate biological mechanisms and clinical relevance. ER, estrogen receptor; HER2, human epidermal growth factor receptor 2. **B,** Scatter plot showing a positive correlation between the *Fidelity Score* and the CNV Score, colored by high- and low-fidelity groups. **C,** Box plots comparing estrogen and HER2 signaling pathway scores between the high- and low-fidelity groups. **D,** Scatter plots showing the correlation between the *Fidelity Score* and the activity of the estrogen signaling (positive) and HER2 signaling (negative) pathways, respectively. Annotated by the seven functional epithelial cell subclusters. **E,** Heatmap showing the interaction weight between the seven functional epithelial cell subclusters and the immune cells. **F,** Spatial visualization of epithelial cell *Fidelity Score* in a representative patient, paired with a hematoxylin and eosin (H&E) staining image. The corresponding box plot compares the *Fidelity Score* between two histological areas. TAM, tumor-associated macrophage. **G,** Violin plots show the number of CAFs, CD8 T cells, TAMs, and endothelial cells adjacent to epithelial cells, stratified by high- or low-fidelity epithelial cell groups. H, *Fidelity Score* in the METABRIC cohort, stratified by molecular subtype and clinical stage. **I,** Kaplan-Meier (K-M) curves demonstrate significantly superior overall survival (OS) for patients in the high *Fidelity Score* group (median value as cutoff) in the METABRIC cohort and for Luminal patients from the METABRIC cohort.

The *Fidelity Score* correlated positively with the CNV score, confirming that high genomic instability may lead to a stronger shedding propensity (**Fig. 6B**). We next investigated the molecular pathways, including estrogen and HER2 signaling, and found that the high-fidelity group was characterized by significantly enriched estrogen receptor (ER) signaling and attenuated HER2 signaling (**Fig. 6C**). Consistent with this, the Fidelity Score correlated with high ER signaling and low HER2 signaling, and each functional epithelial subcluster carried its own pathway signature (**Fig. 6D**). This correlation was further validated in the PDO cohort, with distinct clustering observed across molecular subtypes (Supplementary Fig. S6A). These results suggested that nipple discharge may reveal expression levels of key biomarkers and could therefore help guide treatment decisions.

Cell-cell interaction analysis indicated that the high-fidelity group engaged in stronger interactions with immune cells, suggesting a potential role for the immune microenvironment in regulating the fidelity phenotype (**Fig. 6E**). Ligand-receptor analysis suggested that the downregulated immune interactions in the low-fidelity group were primarily mediated by the MIF-(CD74+CD44) and MIF-(CD74+CXCR4) axes (Supplementary Fig. S6B). Spatial transcriptomics validation further confirmed heterogeneity in the *Fidelity Score* across distinct histological regions (**Fig. 6F**). Moreover, the high-fidelity epithelial group was found to be spatially closer to cancer-associated fibroblasts (CAFs), CD8 T cells, tumor-associated macrophages (TAMs), and endothelial cells than to the low-fidelity group, consistent with the cell–cell interaction analysis (**Fig. 6G**). Single-cell spatial transcriptomics on a slice with well-defined ductal architecture revealed high-fidelity epithelial subclusters localized near the duct lumen (Supplementary Fig. S6C).

In the METABRIC and TCGA-BRCA cohorts, the patient-level Fidelity Score was significantly higher in Luminal (HR-positive) breast cancers than in TNBC and HR-negative/HER2-positive subtypes, suggesting a strong association between shedding propensity and Luminal-type breast cancer. This association was also validated in the PDO cohort (Supplementary Fig. S6D). In the METABRIC and TCGA-BRCA cohorts, the highest Fidelity Score was observed in clinical stage I, indicating that high shedding propensity is an early event in tumor development (**Fig. 6H**; Supplementary Fig. S6E). Most importantly, the Fidelity Score is a robust prognostic indicator. Patients with high Fidelity Scores had significantly better OS than those with low scores in both the METABRIC and TCGA-BRCA cohorts (*p* < 0.0001 and *p* = 0.00032, respectively). This prognostic advantage persisted specifically within Luminal patients across both cohorts (**Fig. 6I**; Supplementary Fig. S6F).

## Discussion

Early diagnosis of cancer is crucial for improving patient prognosis[38], particularly in breast cancer, the most common malignancy in women worldwide[39, 40]. Clinical symptoms remain the foundational element of cancer diagnosis. Nipple discharge is experienced by over 80% of women during their reproductive years[41]. Nevertheless, as one of the principal presenting complaints in breast surgery, its molecular diagnostic potential remains largely unexplored. Therefore, developing early diagnostic methods with high sensitivity and specificity has been a major priority. CNVs are recognized as key early drivers in breast cancer tumorigenesis[42, 43] and serve as promising biomarkers for early-stage detection. Here, we derived a 20-feature CNV panel by analyzing CNV data from TCGA-BRCA (n=1,086). Large-scale validation on GSE87048 (n = 100), GSE118527 (n = 401), METABRIC (n = 2,087), and MSK-CHORD (n = 5,751) confirmed that these CNVs are recurrent, breast-cancer-enriched genomic features. Several implicated genes have been reported as high-frequency driver genes (e.g., the amplified oncogene MYC on AMP10 and the deleted tumor suppressor gene WWOX on DEL05)[43–46].

Integrative single-cell analysis of 50 breast cancer patients across molecular subtypes provided mechanistic insight into this 20-feature CNV panel. Phylogenetic reconstruction showed that 19/20 CNV features were present at early evolutionary nodes in 70% of patients, and 4 features were recurrent in >30% of cases across all molecular subtypes. Notably, the co-occurring AMP07∼12 cluster on chromosomes 8q and 20q was functionally linked to metabolic reprogramming, suggesting that these early-arising amplifications confer bioenergetic advantages that are conserved across molecular subtypes. These findings establish that the CNV features comprising our diagnostic panel represent recurrent, early oncogenic drivers rather than late passenger events.

Significant progress has been made in the use of blood-based liquid biopsies for cancer[11, 47, 48]. Notably, CNV has been employed as an effective biomarker in multiple cancer studies[49, 50]. However, detecting CNV in plasma cfDNA still faces several challenges, including low early-stage ctDNA abundance[51, 52], release influenced by clinical and biological characteristics[53, 54], and clonal hematopoiesis-associated false positives[52, 55]. In contrast, non-blood liquid biopsy specimens, anatomically adjacent to the tumor, are less affected by dilution effects and clonal hematopoiesis[12]. Low-depth detection of CNV biomarkers has already been successfully implemented in non-blood fluids such as urine (for bladder cancer) and cerebrospinal fluid (for central nervous system tumors)[14, 56].

Here, we report a key finding: CNV profiling of nipple discharge yields superior diagnostic performance compared with plasma cfDNA. In the prospective nipple discharge cohort (39 malignant, 18 benign), the panel achieved an AUC of 0.863, with 94.4% specificity and 96.7% PPV, correctly identifying 29/39 cancers while yielding only 1 false-positive. Meanwhile, CNV signals were virtually undetectable in matched plasma cfDNA despite a 10-fold higher sequencing depth. This comparison highlights ndDNA as a locally enriched, tumor-specific DNA source that serves as a high-confidence “rule-in” test to reduce unnecessary invasive diagnostic procedures. Moreover, detection of ndDNA CNVs indicates tumor invasion into the mammary ductal system, suggesting its potential utility in guiding surgical margins.

Compared with blood-based liquid biopsy, our ndDNA strategy offers distinct practical and biological advantages. In GRAIL’s Circulating Cell-free Genome Atlas study of 4,077 participants, a targeted methylation panel at 100× depth achieved 99.5% specificity; however, overall detection sensitivity was 51.5%, and only 16.8% for stage I cancer[57, 58]. The subsequent PATHFINDER prospective study of 6,621 participants confirmed these metrics but still demonstrated low sensitivity for stage I disease[59]. Our model achieved a 70.0% positive rate (14/20) in stage 0 (n = 12) and stage I (n = 8) patients within the *nipple discharge* cohort, indicating strong potential for early detection. Remarkably, while DELFI’s blood-based fragmentomics approach reported a sensitivity of only 57% for breast cancer at 1–2× depth under a comparable 98% specificity threshold[60], our ndDNA framework preserved a substantially higher sensitivity of 74.4% at an ultra-low sequencing depth of ∼1× (requiring only 4 Gb of data). This sharp contrast underscores the biological superiority of tumor-proximal biofluids, which capture concentrated, high-fidelity diagnostic signals that are otherwise heavily diluted by the systemic circulation. Furthermore, blood-based assays typically require 10–20 mL of blood, whereas we obtain equivalent genomic information from only 0.3–0.5 mL of nipple discharge, improving patient adherence and enabling non-invasive self-collection. Prospective integration of ndDNA with imaging should quantify further reductions in invasive downstream procedures.

To elucidate the biological mechanisms underlying this diagnostic efficacy, we used single-cell multi-omics data to trace CNV features to the malignant epithelium, uncovering unequal contributions of different tumor subclusters to ndDNA. We identified a “high-fidelity” group of four epithelial subclusters (Cytoskeleton, Mesenchymal, Mammary Gland, Stress) that faithfully release their CNV features into nipple discharge. Spatial transcriptomic analysis indicated that high-fidelity tumor cells were located closer to the mammary ducts. Further functional analysis revealed a significant correlation between high shedding propensity and high HR signaling and low HER2 signaling. Meanwhile, Luminal cases in TCGA also exhibited higher *Fidelity Scores*, indicating that ndDNA may be more reliable in patients with Luminal breast cancer. Furthermore, the high-fidelity cells displayed significantly stronger interactions with CD8 T cells and TAMs, suggesting that immune-microenvironment cross-talk may facilitate DNA shedding—a hypothesis warranting future validation.

Clinically, high *Fidelity Scores* strongly predicted better outcomes, suggesting that abundant CNV shedding is a marker of a less aggressive, well-differentiated Luminal tumor. Because the score is derived from standard RNA-seq of resected or biopsied tissue, it could be incorporated into existing molecular pathology workflows for early risk stratification. Prospective studies are needed to determine whether high-fidelity tumors also show better response to endocrine or immune-based therapies.

Our study has several limitations. First, the sample size of *the nipple discharge cohort*, while sufficient to demonstrate robust diagnostic accuracy in our first-in-field analysis, will need to be expanded in future multicenter studies to validate and refine the performance of the CNV panel. Because only about 4% of breast cancer patients present with nipple discharge[61], we generalized our findings by validating the CNV panel in two large, independent data sets (TCGA and METABRIC). Nevertheless, even larger prospective cohorts are still required to confirm all observations.

## Conclusions

This work introduces nipple discharge as a highly effective liquid biopsy source for breast cancer diagnosis through CNV profiling. By integrating bulk genomic analyses with single-cell and spatial resolution, we validated a sensitive diagnostic tool, provided a deep biological rationale for its efficacy, and linked diagnostic signal strength to specific tumor cell subpopulations and their underlying biology. These findings pave the way for the clinical translation of nipple discharge-based testing and offer new insights into tumor cell shedding as a biologically and prognostically meaningful phenomenon.

## Declarations

### Ethics approval and consent to participate

This study was approved by the Institutional Review Board (IRB) and the Ethics Committee of the National Cancer Center/National Clinical Research Center for Cancer/Cancer Hospital, Chinese Academy of Medical Sciences and Peking Union Medical College (Approval No. 22/291-3493) and the Affiliated Yantai Yuhuangding Hospital of Qingdao University (Approval No. 2020-289). The research was conducted in accordance with the Declaration of Helsinki. Written informed consent to participate in this study was obtained from all individual participants.

### Consent for publication

Not applicable. The manuscript contains no individual person’s data in any form; all data are presented in aggregated and anonymized form.

### Availability of data and material

The data of *the nipple discharge cohort*, *the plasma cfDNA cohort*, the spatial transcriptomics validation, and *the PDO cohort* are available upon request from the corresponding author, Jiaqi Liu. TCGA-BRCA data are publicly available from the National Cancer Institute Genomic Data Commons portal (https://portal.gdc.cancer.gov). The METABRIC and MSK-CHORD datasets are publicly available on cBioPortal (https://www.cbioportal.org). Data from four validation breast cancer datasets are publicly available from the GEO under accession numbers GSE87048, GSE118527, GSE161529, and GSE199219 (https://www.ncbi.nlm.nih.gov/geo/).

### Author’s contributions

**Y. Li:** Conceptualization, investigation, resources, supervision, data curation, writing–review and editing. **P. Pu:** Conceptualization, formal analysis, visualization, funding acquisition, writing–original draft. **H. Xu:** Software, formal analysis, funding acquisition, methodology, writing–original draft. **R. Zhou:** Investigation, writing–review and editing. **P. Tang:** Investigation, validation, writing–original draft. **W. Wen:** Methodology, supervision, writing–review and editing. **G. Qiao:** Supervision, writing–review and editing. **Y. Zhou:** Resources. **G. Gao:** Resources. **Y. Liu:** Software, formal analysis. **Y. Huang:** Visualization, methodology. **Z. Guo:** Resources, data curation. **T. Shang:** Validation, investigation. **L. Cong:** Validation, investigation. **J. Xiang:** Data curation, visualization. **J. Wu:** Conceptualization, supervision, writing–review and editing. **D. Ma:** Conceptualization, supervision, writing–review and editing. **H. Fang:** Supervision, project administration, writing–review and editing. **J. Liu:** Conceptualization, funding acquisition, supervision, project administration, writing–review and editing. All authors read and approved the final manuscript.

## Acknowledgements

We thank the patients and their families for their participation and for generously providing the nipple discharge samples that made this study possible.

## Supplementary information

**Supplementary Fig. S1.** Identification and validation of copy number variation (CNV) features in breast cancer.

**Supplementary Fig. S2.** Validation of the CNV features diagnostic model in the Molecular Taxonomy of Breast Cancer International Consortium (METABRIC) cohort.

**Supplementary Fig. S3.** Ancestral CNV states in the combined cohort.

**Supplementary Fig. S4.** CNV landscape in nipple discharge samples.

**Supplementary Fig. S5.** Integration of CNV profiles across multiple samples.

**Supplementary Fig. S6.** Additional validation of the functional characterization of high-fidelity epithelial cell groups.

**Supplementary Table 1.** Clinical characteristics of the nipple discharge cohort.

**Supplementary Table 2.** Clinical characteristics of the plasma cfDNA cohort.

**Supplementary Table 3.** Clinical characteristics of the combined cohort.

**Supplementary Table 4.** Details of the 20 breast cancer-specific CNV features selected for the diagnostic panel.

**Supplementary Table 5.** List of genes constituting the Fidelity Score.

**Supplementary File 1.** The inferCNV-based phylogenetic trees for all fifty patients in the combined cohort.

