## Supplementary Figures for "Non-Blood Liquid Biopsy of Nipple Discharge to Decode Tumor-Derived CNV Signatures for Breast Cancer Detection and Prognostication"

**Supplementary Fig. S1. Identification and validation of copy-number variation (CNV) features in breast cancer.**

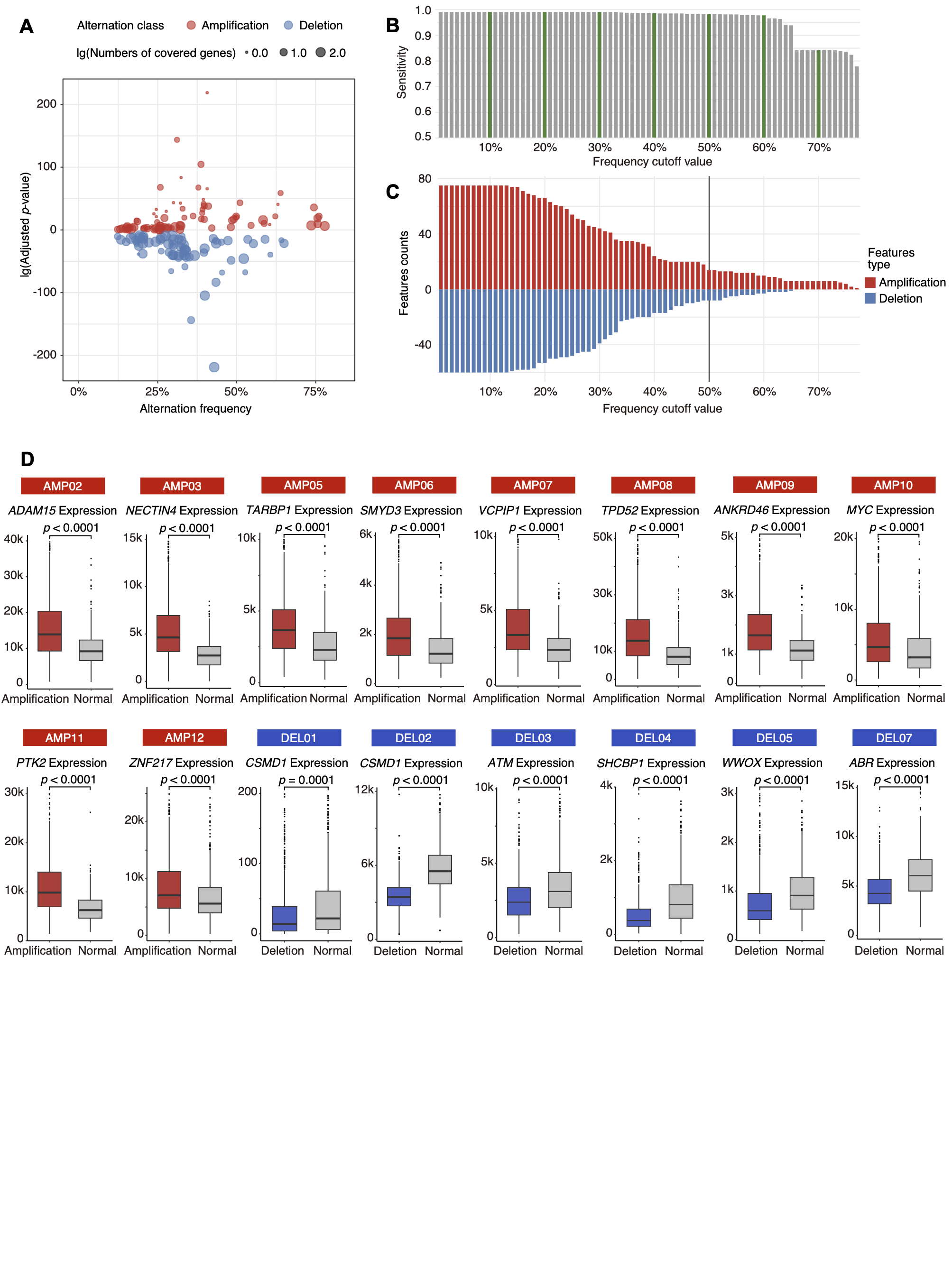

**a,** Dotplot showing the alteration frequency and the -lg(Adjusted p-value) of CNV features identified by the Genomic Identification of Significant Targets in Cancer 2.0 (GISTIC 2.0, v2.0.23) workflow, grouped by alteration class. The dot size corresponds to the lg(Number of covered genes). **b,** Bar plot showing the sensitivity achieved across a range of frequency cutoff thresholds. **c,** Bar plot showing the number of retained amplification and deletion features across a range of frequency cutoff thresholds. **d,** Box plots comparing the expression of representative genes within the CNV features, stratified by alteration state.

**Supplementary Fig. S2. Validation of the CNV features diagnostic model in the Molecular Taxonomy of Breast Cancer International Consortium (METABRIC) cohort.**

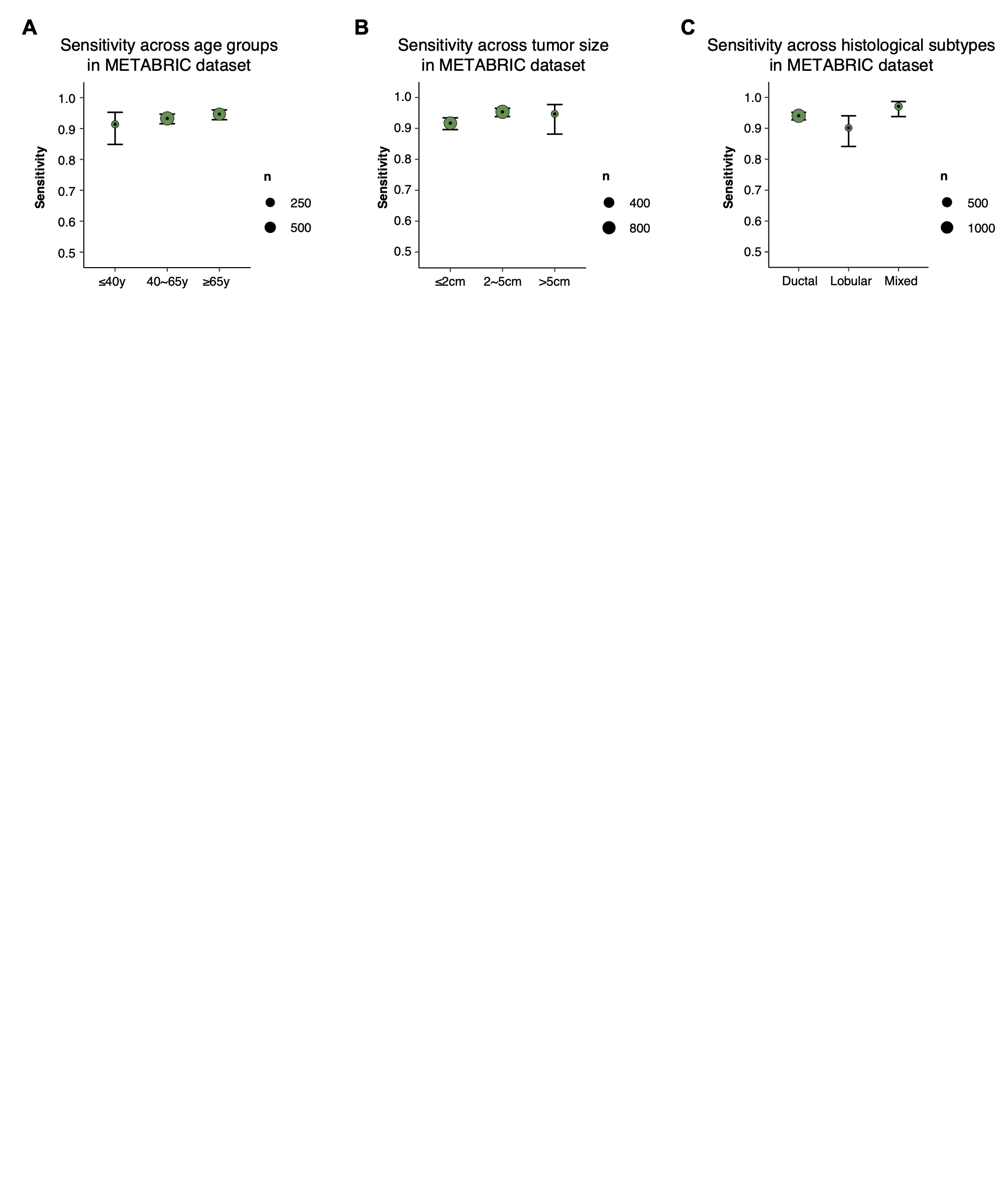

**a-c,** Sensitivity of the CNV features diagnostic model across patient subgroups in the METABRIC cohort, stratified by age (**a**), tumor size (**b**), and histological subtype (**c**). Point estimates (dots) are shown with 95% confidence intervals (error bars). The size of each dot corresponds to the number of patients in the subgroup.

**Supplementary Fig. S3. Ancestral CNV states in the combined cohort.**

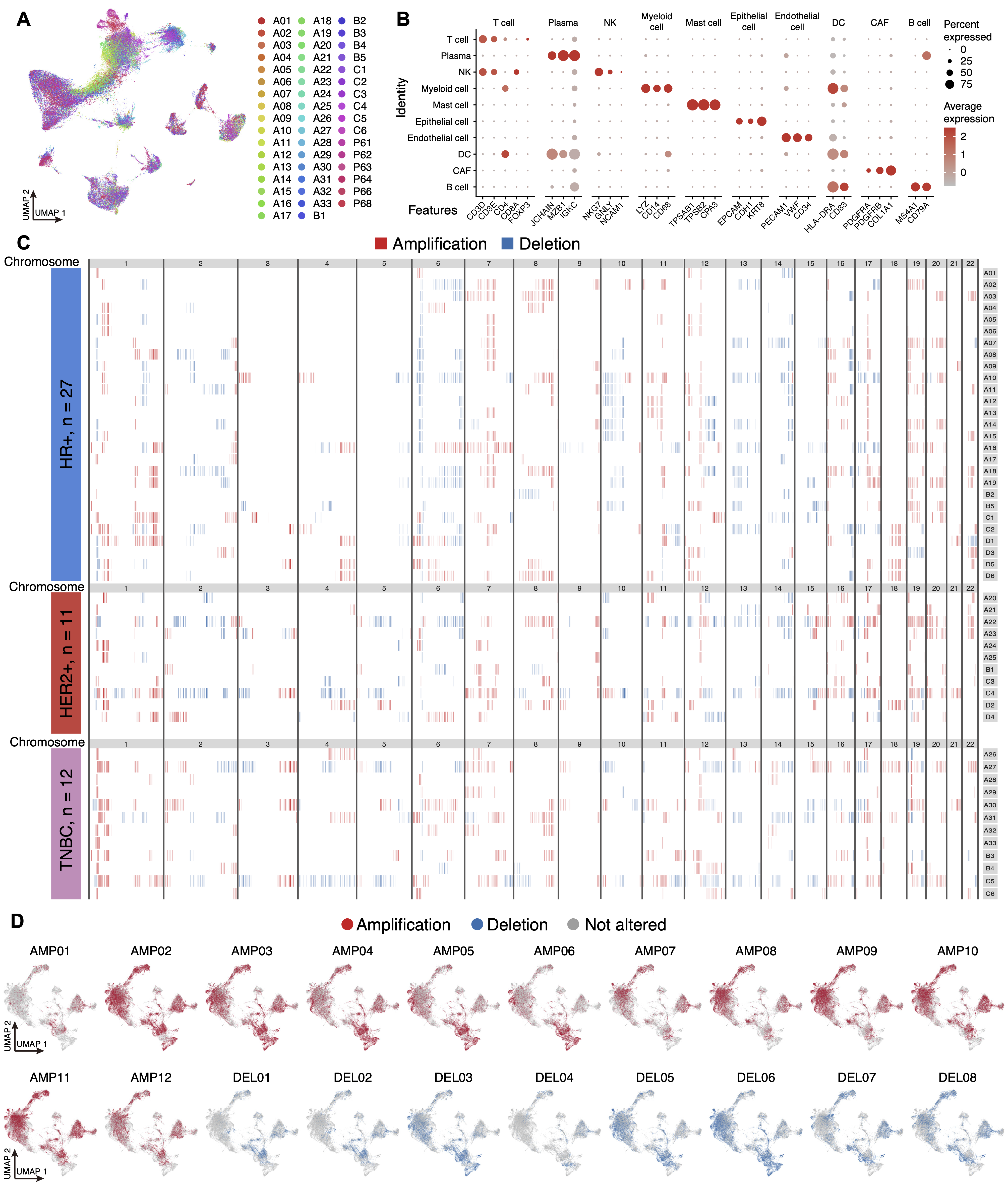

**a,** Uniform Manifold Approximation and Projection (UMAP) visualization of integrated single-cell data from the combined cohort after batch effect correction. **b,** Dot plot showing expression levels of marker genes (x-axis) across cell types (y-axis). **c,** Heatmap showing the genome-wide ancestral CNV state landscape for each patient in the combined cohort. **d,** UMAP of the inferCNV states of the 20 CNV features in epithelial cells from the combined cohort.

**Supplementary Fig. S4. CNV landscape in nipple discharge samples.**

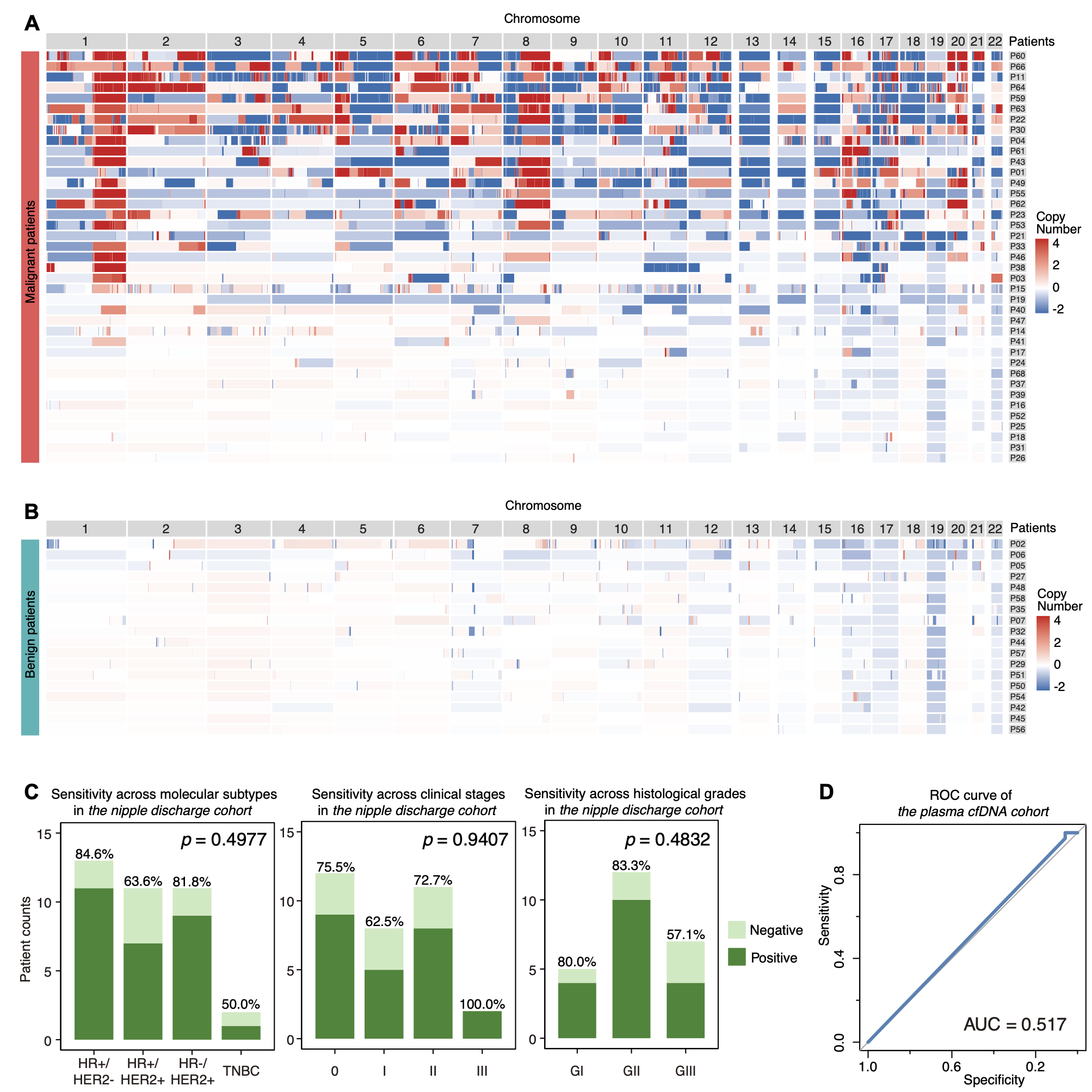

**a,** Heatmap of the genome-wide CNVs landscape for each malignant patient in the nipple discharge cohort. **b,** Heatmap of the genome-wide CNVs landscape for each benign patient in the nipple discharge cohort. **c,** Patient counts and diagnostic sensitivity of the CNV features diagnostic model across patient subgroups in the nipple discharge cohort, stratified by molecular subtype, clinical stage, and histological grade. Fisher's exact test was used to determine *p*-values. **d,** Receiver operator characteristic (ROC) curve for the plasma cfDNA cohort; area under the ROC curve (AUC) = 0.517 (95% CI 0.474–0.560).

**Supplementary Fig. S5. Integration of CNV profiles across multiple samples.**

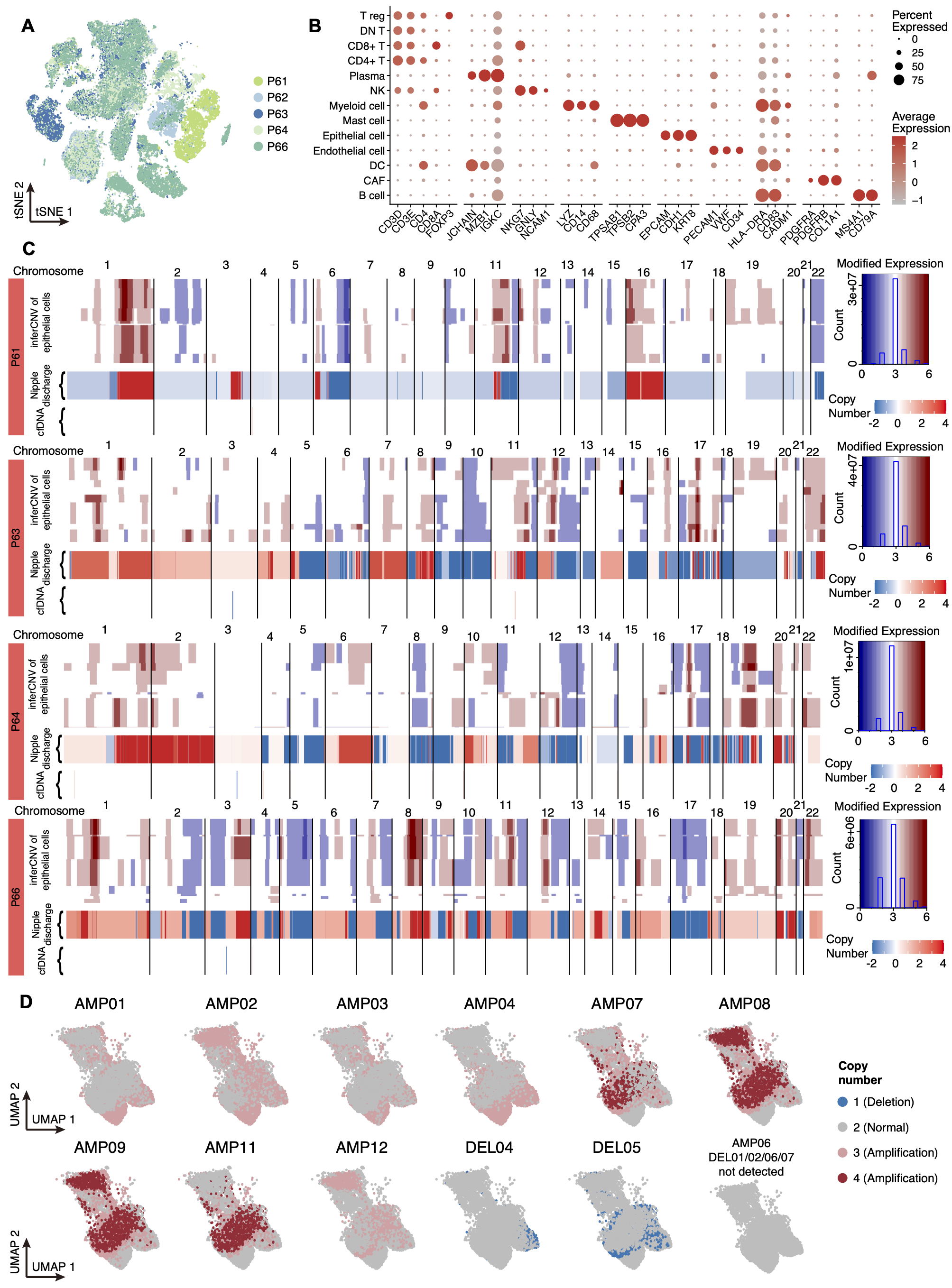

**a,** t-distributed Stochastic Neighbor Embedding (t-SNE) visualization of integrated single-cell data from five patients after batch effect correction. **b,** Dot plot showing expression levels of marker genes (x-axis) across cell types (y-axis). **c,** Genome-wide heatmaps comparing the inferCNV profile of epithelial subclusters with CNV profiles derived from nipple discharge and cfDNA samples in P61, P63, P64, and P66. **d,** UMAP of copy-number states for the remaining CNV features.

**Supplementary Fig. S6. Additional validation of the functional characterization of high-fidelity epithelial cell groups.**

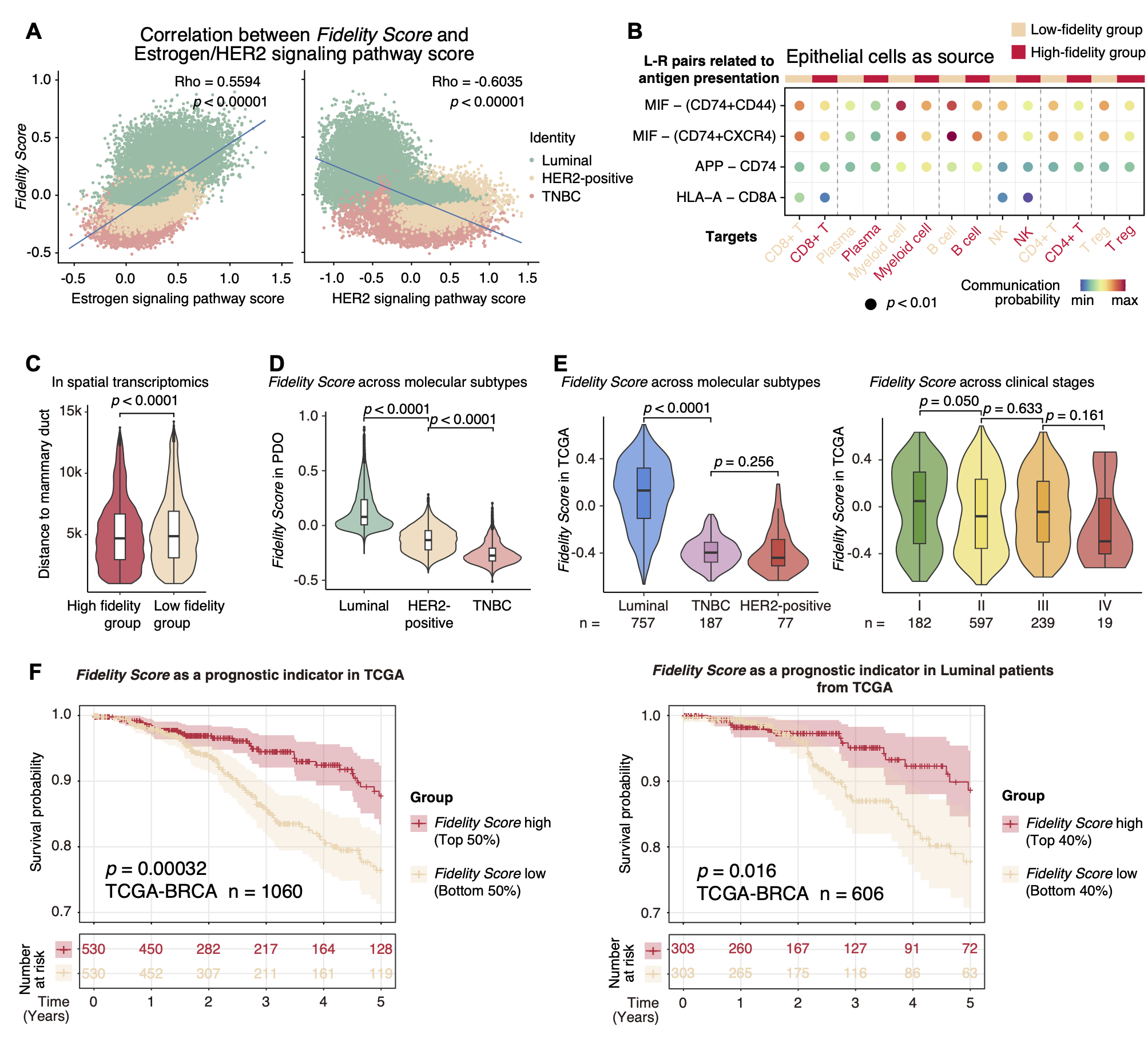

**a,** Scatter plots showing the correlation between the *Fidelity Score* and activity of the estrogen signaling (positive) and HER2 signaling (negative) pathways in the patient-derived organoid (PDO) cohort. **b,** Dot plot showing the communication probability of ligand-receptor pairs associated with immune adjustment from epithelial cells (source) to various immune cell targets, comparing high- versus low-fidelity groups. **c,** Distance to the mammary duct, stratified by high- versus low-fidelity epithelial cell groups. **d,** *Fidelity Score* in the PDO cohort, stratified by molecular subtype. **e,** *Fidelity Score* in the TCGA-BRCA cohort, stratified by molecular subtype and clinical stage. **f,** Kaplan–Meier analysis of overall survival according to high versus low *Fidelity Score*. Left, all TCGA breast cancers (n = 1,060); right, Luminal subset (n = 606).
